# Medical Hallucination in Foundation Models and Their Impact on Healthcare

**DOI:** 10.1101/2025.02.28.25323115

**Authors:** Yubin Kim, Hyewon Jeong, Shan Chen, Shuyue Stella Li, Chanwoo Park, Mingyu Lu, Kumail Alhamoud, Jimin Mun, Cristina Grau, Minseok Jung, Rodrigo Gameiro, Lizhou Fan, Eugene Park, Tristan Lin, Joonsik Yoon, Wonjin Yoon, Maarten Sap, Yulia Tsvetkov, Paul Pu Liang, Xuhai Xu, Xin Liu, Chunjong Park, Hyeonhoon Lee, Hae Won Park, Daniel McDuff, Samir Tulebaev, Cynthia Breazeal

## Abstract

*Hallucinations* in foundation models arise from autoregressive training objectives that prioritize token-likelihood optimization over epistemic accuracy, fostering overconfidence and poorly calibrated uncertainty. In clinical settings, where profound knowledge asymmetry exists between AI systems and end-users, undetected misinformation such as fabricated medications, contraindicated drug recommendations, or false imaging interpretations poses direct patient safety risks. We define **medical hallucination** as any model-generated output that is factually incorrect, logically inconsistent, or unsupported by authoritative clinical evidence in ways that could alter clinical decisions. We evaluated 11 foundation models (7 general-purpose, 4 medical-specialized) across seven medical hallucination tasks spanning medical reasoning, and biomedical information retrieval. General-purpose models achieved significantly higher proportions of hallucination-free responses than medical-specialized models (median: 76.6% vs 51.3%; difference = 25.2%, 95% CI: 18.7–31.3%; Mann–Whitney *U* = 27.0, *p* = 0.012, rank-biserial *r* = *−*0.64). Top-performing model such as Gemini-2.5 Pro exceeded 97% accuracy when augmented with chain-of-thought prompting (base: 87.6%), while medical-specialized models like MedGemma ranged from 28.6–61.9% despite explicit training on medical corpora. Chain-of-thought reasoning significantly reduced hallucinations in 86.4% of tested comparisons after FDR correction (*q <* 0.05), demonstrating that explicit reasoning traces enable self-verification and error detection. Physician audits confirmed that 64–72% of residual hallucinations stemmed from causal or temporal reasoning failures rather than knowledge gaps. A global survey of clinicians (*n* = 70; 15 specialties) validated real-world impact: 91.8% had encountered medical hallucinations, and 84.7% considered them capable of causing patient harm. Our findings reveal medical hallucination as a reasoning-driven failure mode rather than a knowledge deficit. The underperformance of medical-specialized models despite domain training indicates that safety emerges from sophisticated reasoning capabilities and broad knowledge integration developed during large-scale pretraining, not from narrow optimization. Clinical AI safety will therefore require advancing reasoning transparency and adaptive uncertainty management rather than relying on domain-specific fine-tuning alone.

## 1 Introduction

Foundation models are rapidly transforming healthcare, enabling new possibilities in clinical decision support, medical research, and health-system operations [72, 93, 101, 107, 111, 129, 136, 162, 173]. However, their integration into clinical workflows also brings a number of critical challenges to the forefront. A particularly concerning issue is the phenomenon of *hallucination* or *confabulation*, instances where LLMs generate plausible but factually incorrect or fabricated information [35, 90, 106]. Hallucinations are well documented across domains, including finance [102], legal [53], code generation [12], education [46] and more [102, 177, 222]. Hallucination in medical applications pose particularly serious risks as incorrect dosages of medications, drug interactions, or diagnostic criteria can directly lead to life-threatening outcomes [32, 179].

Analogous to cognitive biases in human clinicians [108, 201], LLMs exhibit systematic, context-dependent reasoning errors. We refer to these domain-specific inaccuracies as medical hallucinations; instances where a model produces incorrect, misleading, or unsupported medical information that could influence clinical judgment or patient outcomes and categorize them accordingly (Table 2). For instance, a LLM might hallucinate patient information, history, or symptoms while generating or summarizing a clinical note [203], producing content that diverges from the source record. This example is similar to the confirmation bias of a physician in which contradictory symptoms are overlooked and eventually lead to inappropriate diagnosis and treatment (Fig. 1). Although the concept of hallucinations in LLM is not new [25], its implications in the medical domain warrant specific attention due to the high stakes involved and the minimal margin of error [190].

**Fig. 1:**
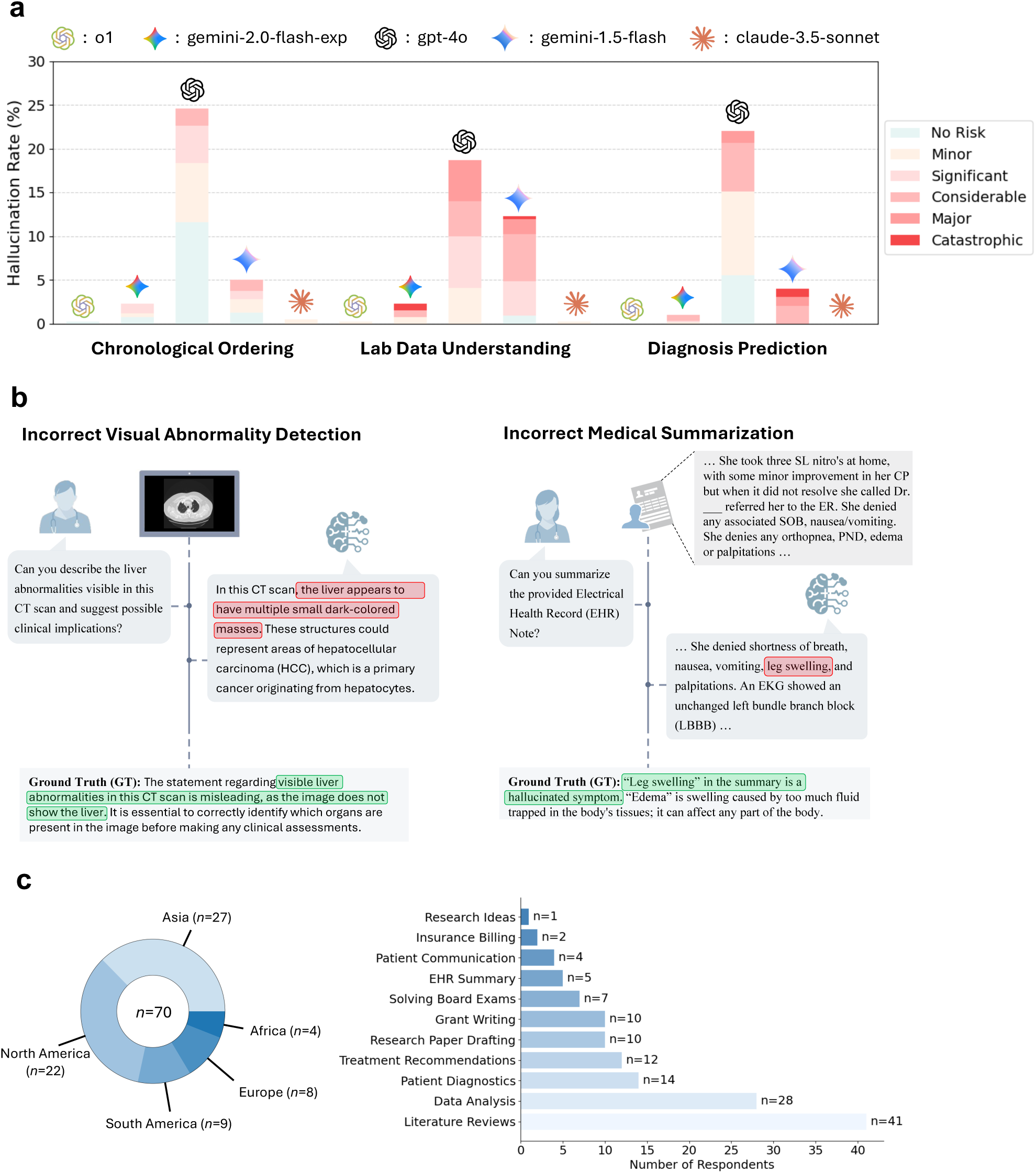
Overview of medical hallucinations generated by state-of-the-art LLMs. (a) Medical expert- annotated hallucination rates and potential risk assessments on three medical reasoning tasks with NEJM Medical Records. The hallucination rate is defined as the percentage of responses containing expert-identified errors (see Section 7 for full analysis). **(b)** Representative examples of medical hallucinations from [44, 203] respectively. **(c)** Geographic distribution of clinician-reported medical hallucination incidents providing a global perspective on the issue (see Subsection 8 for full analysis).

**Fig. 2:**
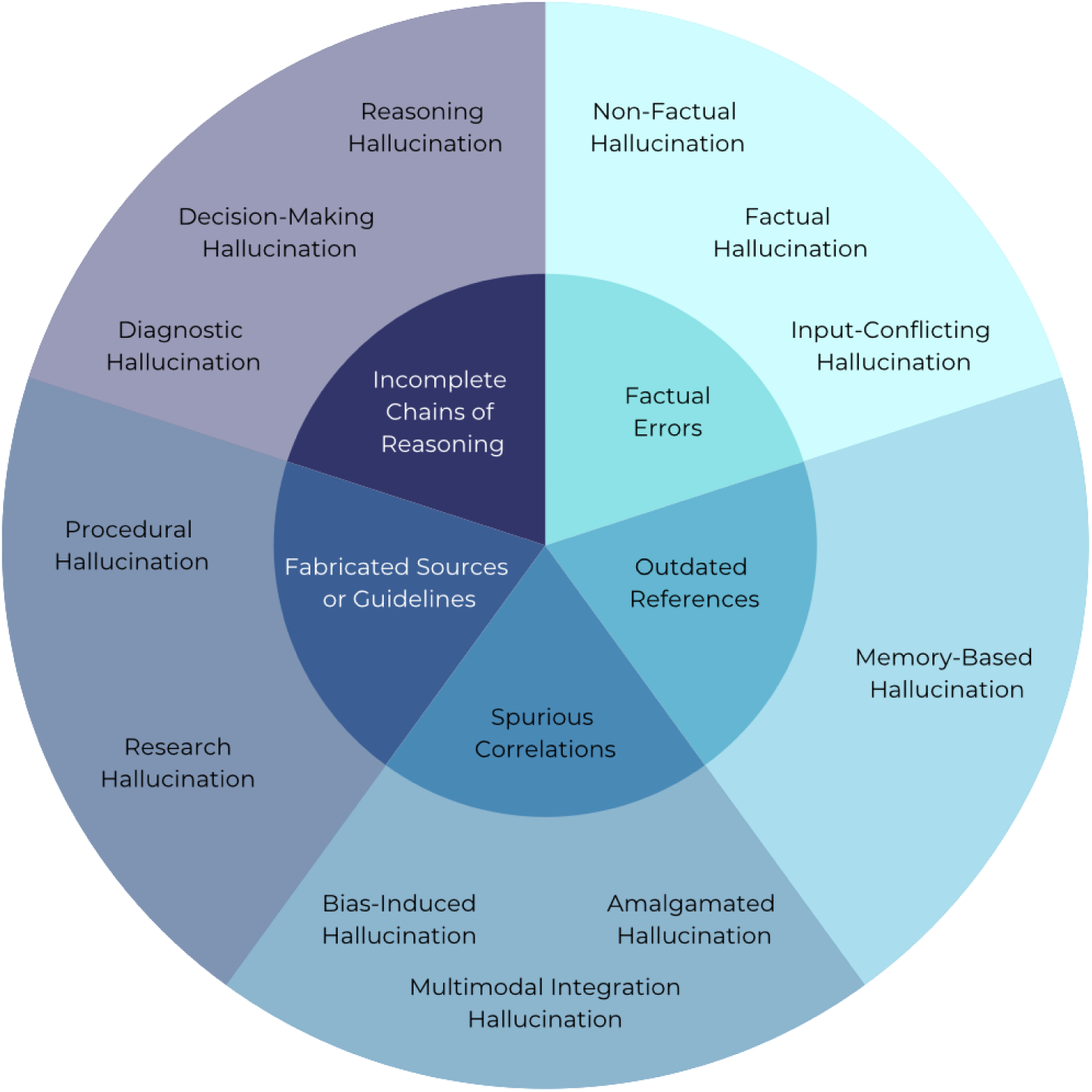
A visual taxonomy of medical hallucinations in LLMs, organized into five main clusters. **(a) Factual Errors**: Hallucinations arising from incorrect or conflicting factual information, encompassing Non- Factual Hallucination, Factual Hallucination, and Input-Conflicting Hallucination. **(b) Outdated References**: Errors due to reliance on outdated or disproven guidelines or data, exemplified by Memory-Based Hallucination. **(c) Spurious Correlations**: Hallucinations that merge or misinterpret data in ways that produce unfounded conclusions, including Bias-Induced Hallucination, Amalgamated Hallucination, and Multimodal Integration Hal- lucination. **(d) Fabricated Sources or Guidelines**: Inventions or misrepresentations of medical procedures and research, covering Procedural Hallucination and Research Hallucination. **(e) Incomplete Chains of Reason- ing**: Flawed or partial logical processes, such as Reasoning Hallucination, Decision-Making Hallucination, and Diagnostic Hallucination.

This paper builds upon existing research on LLM hallucinations [82, 91, 158, 189] and extends it to specific challenges in medical applications, where LLMs face particular hurdles: **1)** the rapid evolution of medical information, leading to potential model obsolescence [216], **2)** the necessity of precision in medical information [186], **3)** the interconnected nature of medical concepts, where a small error can cascade [16], and **4)** the presence of domain-specific jargon and context that require specialized interpretation [237].

Our work makes four primary contributions. **First**, we introduce a taxonomy for medical hallucination in LLMs, providing a structured framework to categorize AI-generated medical misinformation (Table 2). **Second**, we conduct comprehensive experimental analyses across various medical sub-domains including general practice, oncology, cardiology, and medical education, utilizing state-of-the-art LLMs such as GPT-5 [144], Gemini-2.5 Pro [28], and DeepSeek-R1 [50], alongside domain-specific models such as MedGemma [165] (Section 6, Figure 5). **Third**, we present findings from a survey of 70 clinicians, providing empirical insight into how medical professionals perceive and experience hallucinations in their practice or research (Section 8, Figure 9). **Finally**, we explore practical mitigation strategies such as structured prompting and reasoning scaffolds to assess whether these approaches can meaningfully reduce medical hallucination rate across models (Section 6).

**Fig. 3:**
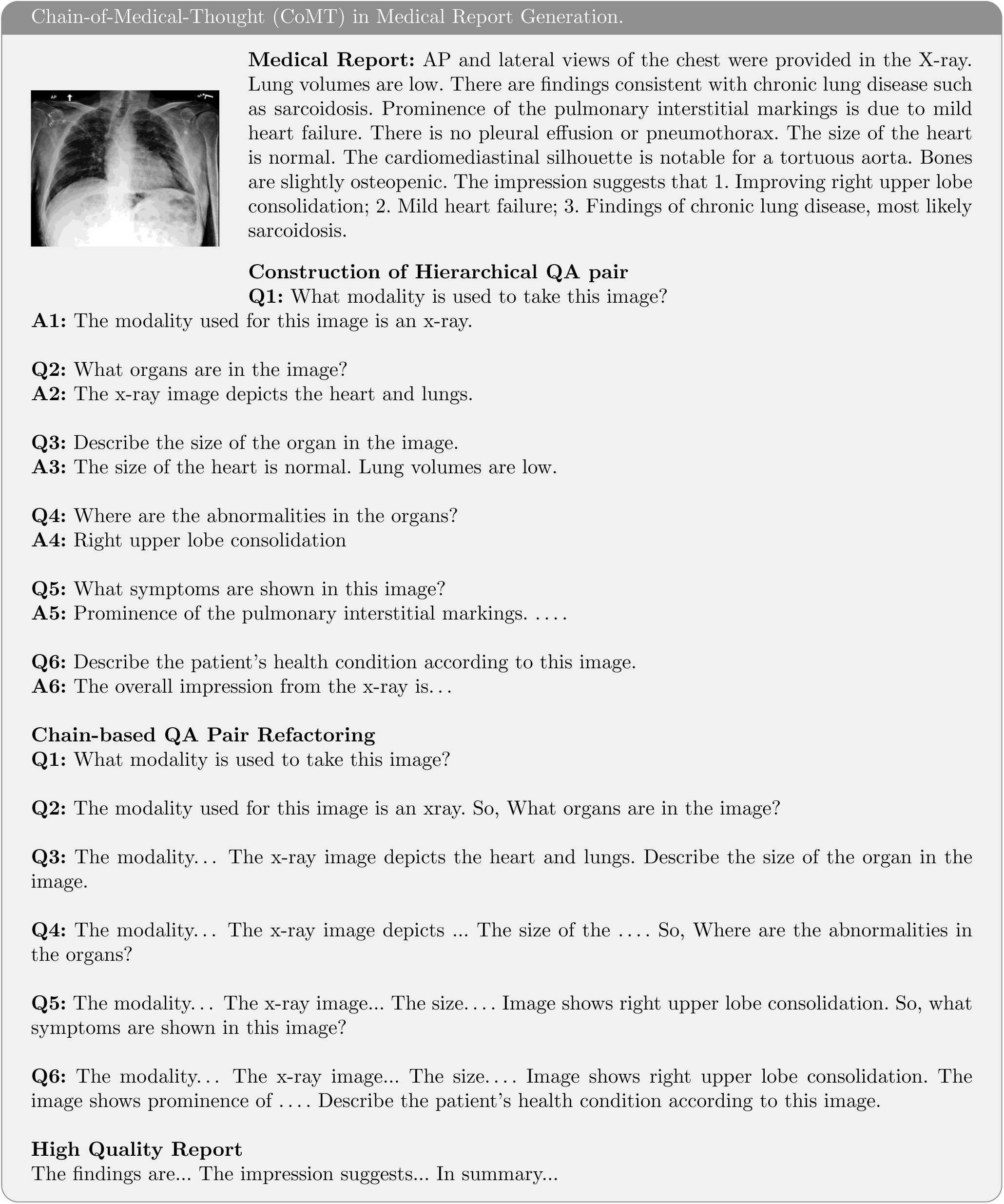
Illustration of CoMT’s process for constructing hierarchical QA pairs based on real clinical image reports. This example is from the original paper [87].

**Fig. 4:**
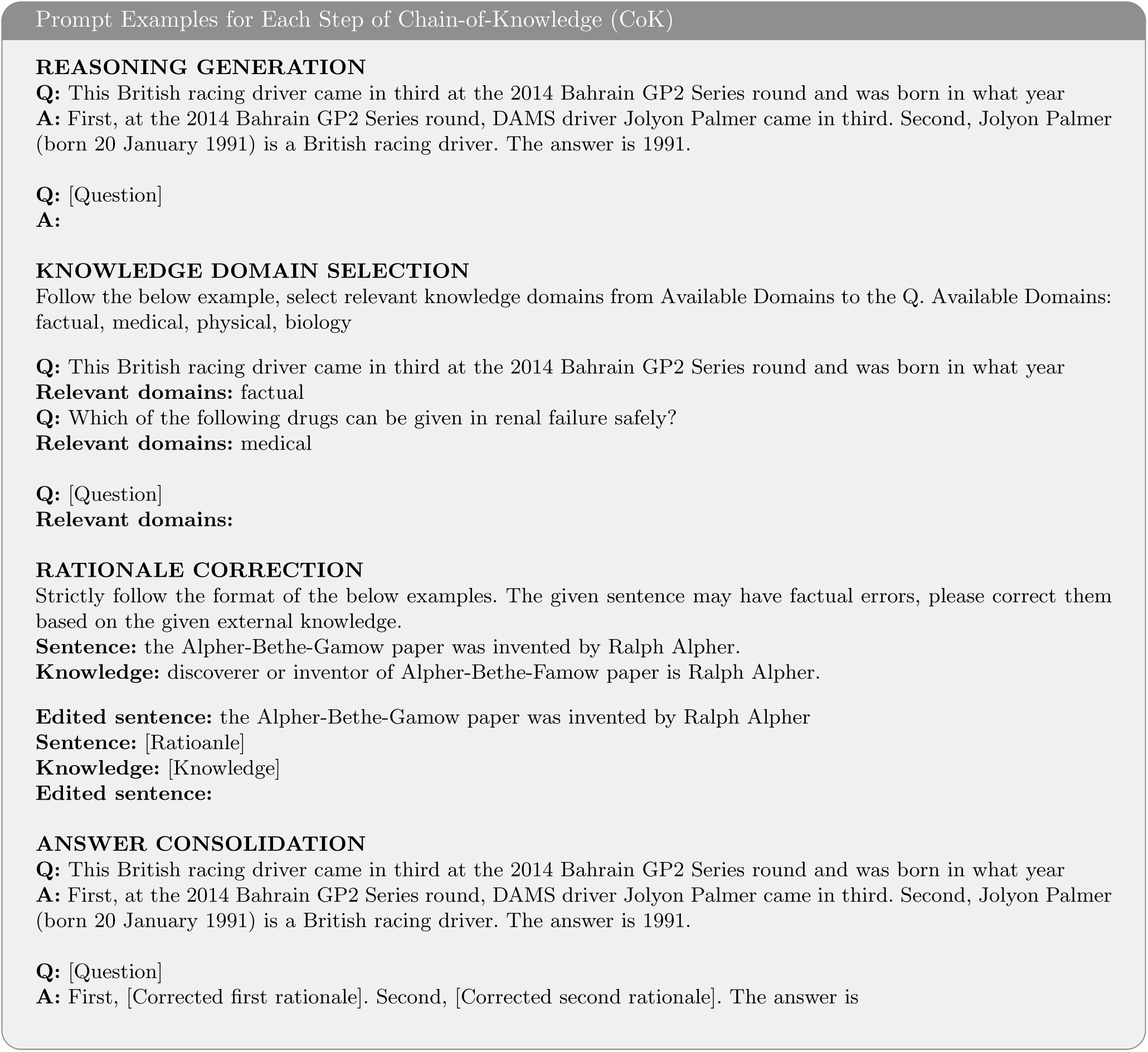
Prompt examples for each step of the Chain-of-Knowledge framework. This example is from the original paper [121].

**Fig. 5:**
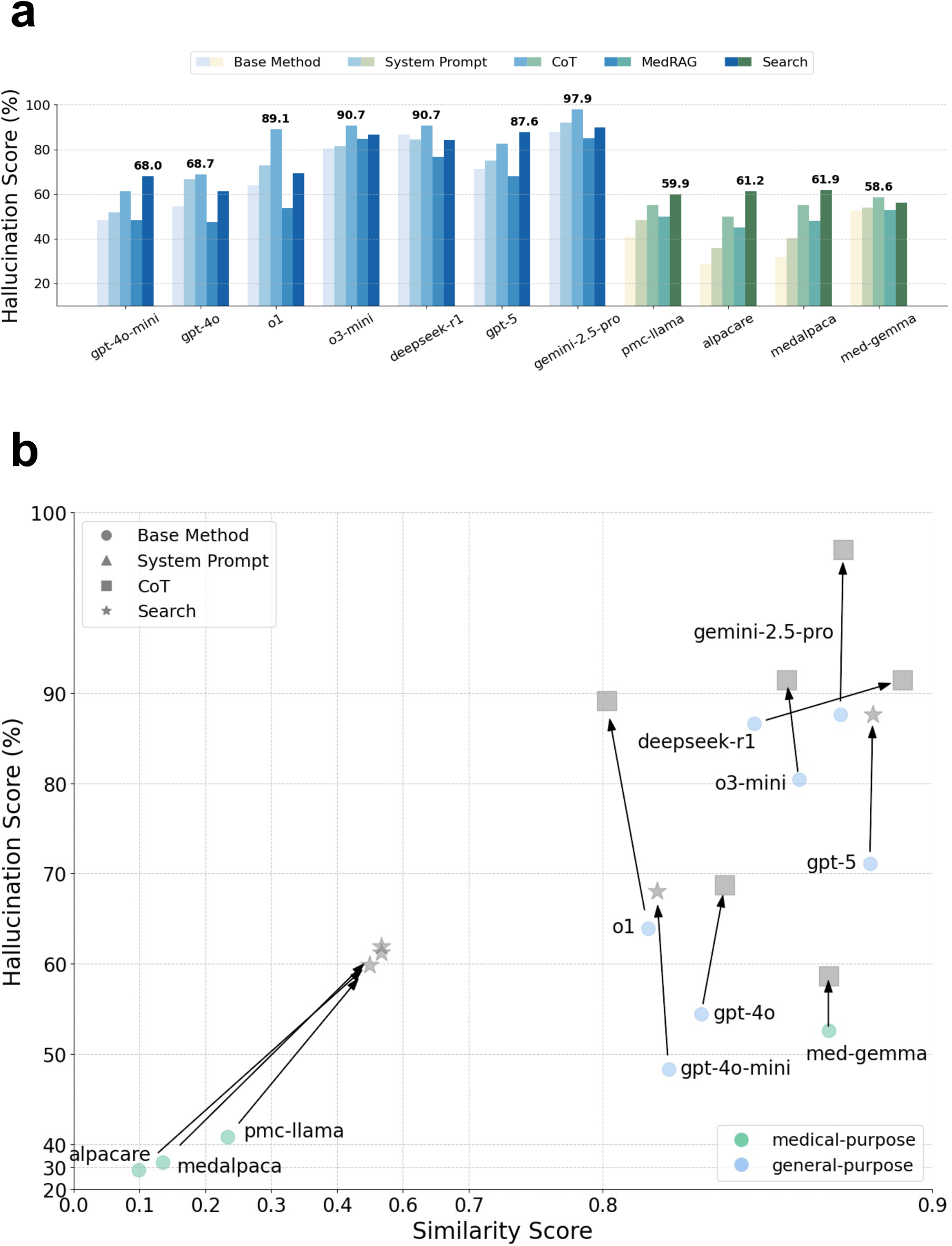
H**a**llucination **Pointwise Score vs. Similarity Score of LLMs on the Med-Halt hallucination benchmark.** This result reveals that the recent advanced models (e.g. o3-mini, deepseek-r1, and gemini-2.5-pro) typically start with high baseline hallucination resistance and tend to see moderate but consistent gains from a simple CoT, while previous models including medical-purpose LLMs often begin at low hallucination resistance yet can benefit from different approaches (e.g. Search, CoT, and System Prompt). Moreover, retrieval-augmented generation can be less effective if the model struggles to reconcile retrieved information with its internal knowledge.

Our findings reveal that even LLMs developed explicitly for medical purposes remain vulnerable to domain-specific hallucinations, often arising from reasoning failures rather than mere knowledge gaps (Section 6). By integrating quantitative benchmarks, physician-led qualitative analysis, and clinician surveys, this work provides the first holistic characterization of medical hallucination in foundation models. These results advance our understanding of AI reliability in medicine and inform regulatory, technical, and ethical frameworks for the safe deployment of clinical AI systems.

## 2 LLM Hallucinations in Medicine

Hallucinations in large language models (LLMs) can undermine the reliability of AI-generated medical information, particularly in clinical settings where inaccuracies may adversely affect patient outcomes. In non-clinical contexts, errors introduced by LLMs may have limited impact or could be more easily detected, particularly because users often possess the background knowledge to verify or cross-reference the information provided, unlike in many medical scenarios where patients may lack the expertise to assess the accuracy of AI-generated medical advice. However, in healthcare, subtle or plausible-sounding misinformation can influence diagnostic reasoning, therapeutic recommendations, or patient counseling [126, 128, 131, 226]. In this section, we define hallucination in a clinical context.

### 2.1 LLMs in Medicine: Capabilities and Adaptations

Recent advancements in transformer-based architectures and large-scale pretraining have elevated LLM performance in tasks requiring language comprehension, contextual reasoning, and multimodal analysis [103, 202]. Examples include OpenAI’s GPT series, Google’s Gemini, Anthropic’s Claude, and Meta’s Llama family. In medicine, researchers adapt these models using domain-specific corpora, instruction tuning, and retrieval-augmented generation (RAG), with the goal of aligning outputs more closely to clinical practice [115, 219].

Several specialized LLMs have demonstrated promising results on medical benchmarks. Med-PaLM and Med-PaLM 2, for instance, exhibit strong performance on tasks such as MedQA [94], MedMCQA [151], and PubMedQA [88] by integrating biomedical texts into their training regimes [161]. Google’s Med-Gemini extends these methods by leveraging multimodal inputs, leading to improved accuracy in clinical evaluations [174]. Open-source initiatives such as MedGemma [165], Meditron [29] and Med42 [37] release models trained on large-scale biomedical data, offering transparency and fostering community-driven improvements^∗^. Despite these tailored efforts, LLMs can generate outputs that appear plausible yet lack factual or logical foundations, manifesting as hallucinations in a clinical context.

A recent survey [141] reinforces these observations, offering a comprehensive review of LLMs in healthcare. In particular, it highlights how domain-specific adaptations such as instruction tuning and retrieval-augmented generation can enhance patient outcomes and streamline medical knowledge dissemination, while also emphasizing the persistent challenges of reliability, interpretability, and hallucination risk.

### 2.2 Differentiating Medical from General Hallucinations

LLM hallucinations refer to outputs that are factually incorrect, logically inconsistent, or inadequately grounded in reliable sources [82]. In general domains, these hallucinations may take the form of factual errors or non-sequiturs. In medicine, they can be more challenging to detect because the language used often appears clinically valid while containing critical inaccuracies [128, 161].

Medical hallucinations exhibit two distinct features compared to their general-purpose counterparts. First, they arise within specialized tasks such as diagnostic reasoning, therapeutic planning, or interpretation of laboratory findings, where inaccuracies have immediate implications for patient care [131, 226, 232]. Second, these hallucinations frequently use domain-specific terms and appear to present coherent logic, which can make them difficult to recognize without expert scrutiny [7, 119]. In settings where clinicians or patients rely on AI recommendations, a tendency potentially heightened in domains like medicine [244], unrecognized errors risk delaying proper interventions or redirecting care pathways.

Moreover, the impact of medical hallucinations is far more severe. Errors in clinical reasoning or misleading treatment recommendations can directly harm patients by delaying proper care or leading to inappropriate interventions [126, 131, 226]. Furthermore, the detectability of such hallucinations depends on the level of domain expertise of the audience and the quality of the prompting provided to the model. Domain experts are more likely to identify subtle inaccuracies in clinical terminology and reasoning, whereas non-experts may struggle to discern these errors, thereby increasing the risk of misinterpretation [7, 119].

These distinctions are crucial: whereas general hallucinations might lead to relatively benign mistakes, medical hallucinations can undermine patient safety and erode trust in AI-assisted clinical systems [7, 119, 126, 131, 152, 226].

### 2.3 Taxonomy of Medical Hallucinations

A growing body of literature proposes frameworks for classifying medical hallucinations in LLM outputs. Agarwal et al. [3] emphasize the severity of errors and their root causes, whereas Ahmad et al. [14] focus on preserving clinician trust by identifying the types of misinformation that most erode confidence. Pal et al. [152] introduce an empirical benchmark for quantifying hallucination frequency in real-world scenarios, underscoring their prevalence. Efforts by Hegselmann et al. [80] and Moradi et al. [124] highlight how data quality and curation practices can influence hallucination rates, especially in the context of patient summaries.

Informed by these studies [7, 112, 113, 146, 207, 234, 246, 249, 250], we first illustrate our taxonomy in Figure 2, which clusters hallucinations into five main categories (factual errors, outdated references, spurious correlations, incomplete chains of reasoning, and fabricated sources or guidelines) based on their underlying causes and manifestations. Subsequently, we present a more granular breakdown of each hallucination types and their exampels in Table 2 and present the various ways in which clinically oriented LLMs may produce superficially plausible but ultimately incorrect outputs.

These hallucinations are exacerbated by the complexity and specificity of medical knowledge, where subtle differences in terminology or reasoning can lead to significant misunderstandings [128, 131]. Furthermore, as shown in Table 1, these hallucinations can manifest across a wide range of medical tasks, from symptom diagnosis and patient management to the interpretation of lab results and visual data.

**Table 1:**
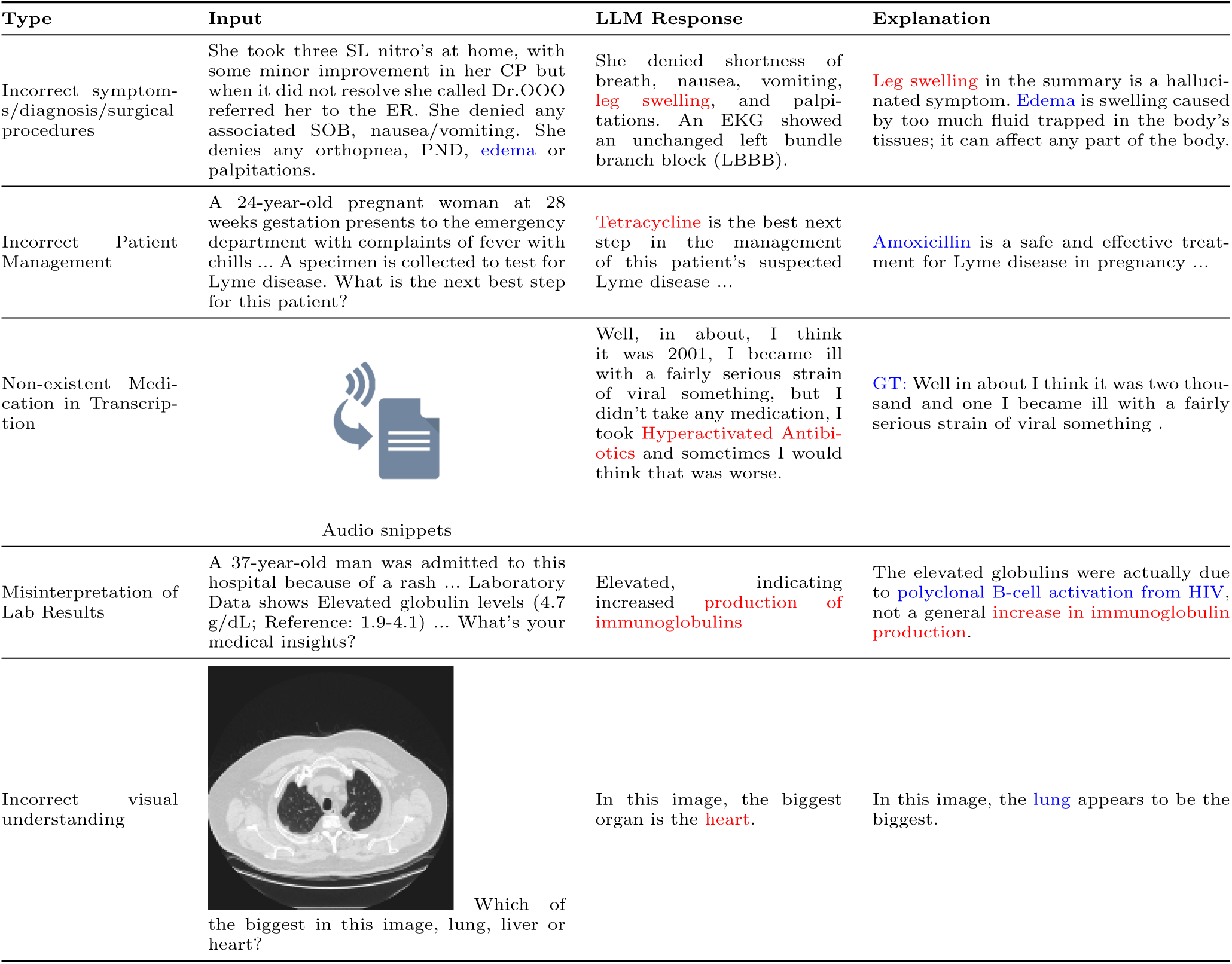
Example types of medical hallucination in clinical tasks. Incorrect or hallucinated information is highlighted in red, while correct explanations are highlighted in blue. These hallucinations span multiple modalities demonstrating the widespread nature of errors in foundation models. These examples are from [21, 22, 44, 152, 203] respectively.

**Table 2:** A taxonomy of medical hallucinations. The table categorizes different types of medical hallucinations, defining each type and providing real-world scenarios along with illustrative examples.

| Type | Definition | Scenario | Example |
| --- | --- | --- | --- |
| Amalgamated Hallucination | Merging unrelated information into a single response, creating factually incoherent outputs. | A patient with type 2 diabetes also has a skin infection. | *“Use insulin topically for 5 days to treat both infection and blood glucose.”* Incorrectly merges insulin therapy with antibiotic ointment, risking confusion and improper care. |
| Non-Factual Hallucination | Fabricating plausible but incorrect information without grounding in factual data. | The LLM is asked about a drug that does not exist in real pharmacopeia. | *“Penmerol is the recommended medication.”* Invents a drug name with no clinical basis, misleading clinicians and undermining trust. |
| Input-Conflicting Hallucination | Generating content that conflicts with the provided input, such as task instructions or task data. | Patient notes indicate a severe penicillin allergy. | *“Prescribe amoxicillin for the infection.”* Overlooks documented allergy, potentially causing severe adverse reactions. |
| Reasoning Hallucination | Failing logical reasoning or problem-solving tasks, leading to flawed clinical recommendations. | The LLM must explain the pathophysiology of congestive heart failure (CHF). | *“CHF is primarily caused by lung infections.”* Provides a logical-sounding but incorrect chain of thought, jeopardizing accurate diagnosis and treatment. |
| Memory-Based Hallucination | Inaccurately recalling or fabricating information the model was trained to retrieve. | The LLM is asked for current hypertension guidelines. | *“According to the 2023 HPC guidelines, start with drug X.”* Quotes a non-existent or outdated protocol, potentially causing suboptimal or incorrect care. |
| Decision-Making Hallucination | Suggesting inappropriate treatment plans or clinical decisions due to flawed reasoning or data interpretation. | A pregnant patient with hypertension consults the LLM for medication guidance. | *“Medication X is safe in pregnancy.”* In reality, it is contraindicated, risking harm to both mother and fetus. |
| Diagnostic Hallucination | Proposing incorrect diagnoses or misinterpreting clinical signs, leading to potential misdiagnosis. | The LLM is given lab results inconsistent with pneumonia. | *“This patient has pneumonia.”* Confidently diagnoses pneumonia despite contradictory evidence, delaying correct treatment and risking patient harm. |
| Procedural Hallucination | Errors in describing medical procedures or protocols. | A user asks for a guide to laparoscopic cholecystectomy. | *“Apply glutaraldehyde to the gallbladder before removal.”* Invents an unrecognized surgical step, causing confusion or potential complications if followed. |
| Factual Hallucination | Generating incorrect factual information, critical in clinical documentation and automation. | A user queries the LLM about the FDA approval status of a newly introduced medication. | *“Drug X is FDA-approved for condition Y.”* Claims an unapproved drug is authorized, potentially prompting off-label or inappropriate use. |
| Bias-Induced Hallucination | Reflecting or amplifying biases in training data, leading to discriminatory or skewed outputs. | The LLM evaluates a minority patient with atypical symptoms. | *“Patients of X background rarely develop condition Y.”* Overlooks key symptoms or leans on stereotypes, perpetuating healthcare disparities. |
| Research Hallucination | Misinterpreting or fabricating research data, potentially impacting evidence-based medicine or drug development. | The LLM is asked about recent clinical trials for a novel anticancer drug. | *“A Phase III trial confirmed 90% efficacy.”* References a non-existent trial, leading clinicians to believe there is strong evidence for efficacy when none exists. |
| Multimodal Integration Hallucination | Errors in interpreting data from multiple sources (e.g., text, imaging, lab results). | The LLM receives both a CT scan image and a textual report indicating a suspicious liver lesion. | *“CT imaging shows no liver abnormalities.”* Contradicts the textual findings, resulting in missed or delayed diagnosis. |

### 2.4 Medical Hallucinations vs. Cognitive Biases: Different Origins, Similar Outcomes

Cognitive biases in medical practice are well-studied phenomena, whereby clinicians deviate from optimal decision-making due to systematic errors in judgment and reasoning [15, 143, 169, 182]. These biases frequently arise in time-constrained or high-stress environments and can undermine the diagnostic and therapeutic process. Although LLMs do not possess human psychology, the erroneous outputs they produce often exhibit patterns that resemble these biases in clinical reasoning. By comparing medical hallucinations in LLMs to established cognitive biases, researchers gain insights into both the roots of AI-driven errors and potential strategies to mitigate them.

#### Common Cognitive Biases in Clinical Practice parallels with LLM Hallucinations

Clinicians frequently experience biases such as *anchoring bias*, which entails relying excessively on initial impressions, even when new evidence suggests alternative explanations. Similarly, *confirmation bias* leads to selective acceptance of data that reinforces a working diagnosis, while *availability bias* skews judgments toward diagnoses that are more memorable or have been recently encountered [15, 78, 118, 157]. Clinicians also exhibit *overconfidence bias*, characterized by unwarranted certainty in diagnostic or therapeutic decisions [126, 169], as well as *premature closure*, where they settle on a plausible explanation without fully considering differential diagnoses [15].

Hallucinations in medical LLMs echo these biases in various ways. *Anchoring* appears when a model disproportionately relies on the initial part of a prompt, neglecting subsequent details or contextual information. *Confirmation bias* emerges when an LLM’s response aligns too closely with the user’s implied hypothesis, neglecting contradictory evidence. *Availability* manifests in the model’s tendency to propose diagnoses or treatments that are disproportionately represented in its training data. *Overconfidence* becomes evident when LLM outputs present an unwarranted level of certainty, a phenomenon linked to poor calibration [43, 74]. Finally, *premature closure* can occur if the model settles on a single, plausible-sounding conclusion without comprehensively considering differential possibilities or additional context [80]. Although the LLM lacks human cognition, the statistical patterns it learns can simulate biases that arise from heuristic-based thinking in clinicians.

Despite these surface-level similarities, the basis of LLM hallucinations diverges from the cognitive underpinnings of human biases. Cognitive biases result from heuristic shortcuts, emotional influences, memory limitations, and other psychological factors [143, 182]. In contrast, medical LLM hallucinations are the product of learned statistical correlations in training data, coupled with architectural constraints such as limited causal reasoning [66, 92]. This distinction means that while clinicians might fail to adjust their thinking in light of conflicting information, an LLM may simply lack exposure to correct or more recent evidence—or fail to retrieve it—leading to erroneous outputs. Consequently, mitigation requires strategies tailored to each context: clinicians might benefit from decision-support tools and reflective practice to counter personal biases, while LLMs demand better data curation, retrieval-augmented generation, or explicit calibration methods to curb hallucinations and unwarranted certainty.

Identifying parallels between cognitive biases and LLM hallucinations highlights potential remediation avenues. Techniques for reducing *anchoring* and *confirmation bias* in clinical settings—such as prompting systematic consideration of differential diagnoses—may inform prompt design or chain-of-thought strategies in LLMs [223]. Encouraging models to output uncertainty estimates or alternative explanations can address *overconfidence* and *premature closure* biases, especially if users are guided to critically evaluate multiple options. Meanwhile, robust fine-tuning procedures and retrieval-augmented generation can improve the balance of training data, mitigating the model’s *availability* bias. Taken together, these efforts could reduce the frequency and severity of hallucinations, ensuring AI-assisted systems more closely align with evidence-based clinical practice.

### 2.5 Clinical Implications of Medical Hallucinations

The integration of large language models (LLMs), which remain susceptible to hallucination, into healthcare introduces significant risks with direct implications for patient safety and clinical practice. Hallucinated outputs that appear credible can guide clinicians toward ineffective or harmful interventions, influencing therapeutic choices, diagnostic pathways, and patient-provider communication [81, 126, 186]. We present how such hallucinations undermine patient safety, disrupt clinical workflows, and create additional ethical and legal complexities.

A chief concern is **patient safety**. When hallucinated outputs lead to incorrect recommendations or misdiagnoses, clinicians may adopt interventions that inadvertently harm patients [81]. Even minor inaccuracies can escalate clinical risks if they go unnoticed or align with a clinician’s cognitive bias, ultimately compromising the quality of care.

Another critical dimension is the **erosion of trust** in AI systems. Repeated hallucinations often breed skepticism among both healthcare providers and patients [204]. Providers are less inclined to rely on potentially error-prone models, while patients may grow apprehensive about the reliability of artificial intelligence in medical decisions, inhibiting broader integration of these tools in clinical practice.

These errors also disrupt **workflow efficiency**. Hallucinations can force clinicians to verify or correct AI-generated information, adding to their workload and diverting attention from direct patient care [127]. This burden can diminish the potential benefits of automation and decision support, particularly in time-sensitive or resource-constrained environments.

Beyond immediate bedside concerns, **ethical and legal implications** arise from the growing reliance on LLM-based recommendations [60]. As models increasingly influence clinical decision-making, the question of accountability for AI-driven errors becomes more urgent. Uncertainty over liability may impede system-wide adoption and complicate the legal landscape for healthcare providers, technology developers, and regulators.

Finally, hallucinations curtail the **impact on precision medicine** by reducing the trustworthiness of personalized treatment recommendations. If an LLM’s outputs cannot consistently deliver accurate, context-specific insights, it undermines the potential to tailor interventions to individual patient profiles [200]. The vision of leveraging big data to refine therapeutic strategies becomes more challenging if model-generated advice contains undetected inaccuracies.

Overall, understanding the clinical implications of medical hallucinations is essential for developing safer, more trustworthy models in healthcare. Stakeholders must consider patient safety, provider engagement, and the broader ethical and legal context to ensure that emerging technologies ultimately enhance, rather than impede, medical practice.

## 3 Causes of Hallucinations

Hallucinations in medical LLMs often arise from a confluence of factors relating to data, model architecture, and the unique complexities of healthcare. In this section, we build up on Section 2 where we presented how hallucinations manifest and why they matter in clinical contexts and provide a deeper look at the root causes. By understanding where and how LLMs fail, researchers and practitioners can prioritize interventions that safeguard patient well-being and advance the reliability of AI in medicine.

### 3.1 Data-Related Factors

#### Data Quality and Noise

Clinical datasets, such as electronic health record (EHR) and physician notes, often contain noise in the form of incomplete entries, misspellings, and ambiguous abbreviations. These inconsistencies propagate errors into LLM training [80]. For instance, a lack of structured input may confuse models, leading them to replicate false patterns or irrelevant outputs [124]. Outdated data further compounds this issue. Medical knowledge evolves continuously, and guidelines can quickly become outdated [167]. Models trained on static or historical data may recommend ineffective treatments, reducing clinical utility [66]. Addressing these issues requires rigorous data curation, including noise filtering, deduplication, and alignment with current medical guidelines.

#### Data Diversity and Representativeness

Training data must reflect the diversity of patient populations, disease presentations, and healthcare systems. Biased datasets, such as those dominated by common conditions or data from high-resource settings, limit model generalizability [31, 42, 96]. For instance, underrepresentation of minority groups can lead to systematic errors in AI predictions. Rare diseases are particularly affected [160]. Models often lack exposure to these conditions during training, leading to hallucinations when generating diagnostic insights [164]. Similarly, regional variations in clinical terminology and disease prevalence further exacerbate performance disparities [141]. Standardized terminologies, such as Systematized Nomenclature of Medicine Clinical Terms (SNOMED CT), can improve consistency by harmonizing medical language across datasets [45]. Efforts to mitigate data diversity challenges include targeted inclusion of underrepresented conditions and populations, as well as benchmarking on globally diverse datasets to assess generalizability [31, 68, 125].

#### Size and Scope of Training Data

While large-scale datasets are critical for training LLMs, general-purpose models often lack sufficient exposure to domain-specific medical content. As demonstrated by Alsentzer et al. [6], fine-tuning models on biomedical corpora significantly improves their understanding of clinical text. In contrast, inadequate training data coverage creates knowledge gaps, causing models to hallucinate when addressing unfamiliar medical topics [116]. Comprehensive training datasets that incorporate annotated clinical notes, peer-reviewed research, and real-world guidelines are essential to ensure coverage of both common and edge cases. Expanding the scope to include rare diseases, specialized treatments, and emerging conditions can further enhance model reliability [92].

#### Ambiguity in Clinical Language

Unique complexities in the medical domain exacerbate hallucinations in LLMs, such as ambiguity in clinical language, or rapidly evolving nature of medical knowledge. Medical text often contains *ambiguous abbreviations*, incomplete sentences, and inconsistent terminology. For example, “BP” could mean “blood pressure” or “biopsy,” depending on context [80]. Such ambiguities challenge LLMs, leading to misinterpretations and hallucinations. Standardization efforts, such as the adoption of structured vocabularies like SNOMED CT [45] provide consistent mappings of clinical terminology, reducing ambiguity and improving model reliability.

#### Rapidly Evolving Medical Knowledge

Furthermore, medical knowledge evolves continuously as new treatments, guidelines, and evidence emerge. Static training datasets quickly become outdated, causing models to generate recommendations that no longer reflect current clinical best practices [160, 167]. To address this, models require regular fine-tuning on updated medical data and integration with dynamic knowledge retrieval systems. Tools capable of real-time evidence synthesis can help ensure outputs remain clinically relevant [26, 141].

### 3.2 Model-Related Factors

Limitations intrinsic to model architecture and behavior also contribute to medical hallucinations, independently or in combination with clinical data related factors introduced in Section 3.1.

#### Overconfidence and Calibration

LLMs frequently exhibit overconfidence, generating outputs with high certainty even when the information is incorrect. Poor calibration—where confidence scores fail to align with prediction accuracy—can mislead clinicians into trusting inaccurate outputs [43]. For example, Yuan et al. [242] highlight the need for improved uncertainty estimation techniques to mitigate overconfidence. Effective strategies for addressing calibration include probabilistic modeling, confidence-aware training, and ensemble methods. These approaches enable models to provide uncertainty estimates alongside predictions, promoting safer integration into clinical workflows.

#### Generalization to Unseen Cases

Medical LLMs struggle to generalize beyond their training data, particularly when faced with rare diseases, novel treatments, or atypical clinical presentations. Models trained on imbalanced datasets often extrapolate from unrelated patterns, producing erroneous or irrelevant outputs [80, 164]. Wang et al. [206] demonstrate that general-purpose LLMs require domain-specific fine-tuning to adapt effectively to clinical tasks. Additionally, retrieval-augmented generation (RAG) techniques, which allow models to access external knowledge dynamically, can help improve performance on unfamiliar cases [116].

#### Lack of Medical Reasoning

Effective clinical decision-making relies on a complex cognitive process known as medical reasoning, which involves integrating patient symptoms, medical history, diagnostic data, and evidence-based treatments to formulate a differential diagnosis and treatment plan. It is a step beyond simple information retrieval, requiring causal inference and contextual understanding. However, LLMs primarily rely on statistical correlations learned from text rather than true causal reasoning [92]. As a result, hallucinations occur when models generate outputs that sound plausible but lack logical coherence [66]. For example, an LLM might correctly identify that chest pain is a symptom of a heart attack but fail to reason that in a 20-year-old patient with no risk factors, this symptom is more likely indicative of musculoskeletal pain or anxiety. This failure to weigh probabilities and consider the full clinical context is a hallmark of a breakdown in medical reasoning. Structured knowledge integration, such as incorporating clinical pathways and causal frameworks into training, has shown potential in improving model reasoning. Similarly, prompting strategies, such as CoT reasoning, can encourage step-by-step output generation to better mimic clinical thought processes [223].

## 4 Detection and Evaluation of Medical Hallucinations

### 4.1 Hallucination Detection Methods

We explore several general strategies for hallucination detection in LLMs, alongside some healthcare-specific approaches. Hallucinations in LLMs occur when the model generates outputs that are unsupported by factual knowledge or the input context. Detection methods can be broadly categorized into three groups: 1) factual verification, 2) summary consistency verification, and 3) uncertainty-based hallucination detection. Various benchmarks have been developed to evaluate the effectiveness of these detection strategies, as summarized in Table 3.

**Table 3:** An overview of medical hallucination benchmarks. The table summarizes existing benchmarks designed to evaluate hallucinations in medical contexts, showcasing the diversity of task types, input data sources, and evaluation metrics. These benchmarks span multiple domains, including medical exams, radiology reports, clinical histories, and LLM-generated summaries, with evaluation criteria ranging from accuracy and confidence scoring to fluency, coherence, and error reduction.

| Benchmark | Task Type | Input | Metric |
| --- | --- | --- | --- |
| Med-HALT<br>[152] | 1) False Confidence Test<br>2) None of the Above Test<br>3) Fake Question Test<br>4) Memory Hallucination Tests | Medical exams<br>PubMed | Pointwise Score<br>Accuracy |
| HALT-MedVQA<br>[208] | 1) FAKE Question<br>2) None of the Above<br>3) Image SWAP | Visual Medical Query | Accuracy |
| CMHE-HD<br>[55] | 1) Hallucination Detection<br>2) Disease Diagnosis<br>3) Concept Explaining | Clinical histories<br>EHR<br>Medical Conversations | Accuracy |
| Med-VH<br>[71] | 1) Wrongful Image<br>2) False Confidence Justification<br>3) None of the Above<br>4) Report Generation<br>5) Clinically Incorrect Premise | Chest X-rays<br>Clinical Reports<br>Diagnostic Questions | Knowledge Accuracy<br>Hallucinated Rate<br>Capacitation Score<br>CHAIR Score |
| Med-HallMark<br>[44] | 1) Catastrophic Hallucination<br>2) Critical Hallucination<br>3) Attribute Hallucination<br>4) Prompt-induced Hallucination<br>5) Minor Hallucination | Medical Exams<br>Chest X-rays<br>CT Scans<br>MRI Images<br>Radiology Reports | BertScore<br>METEOR<br>ROUGE<br>BLEU<br>MediHall Score<br>Accuracy |
| K-QA<br>[134] | 1) Long-form answer annotation<br>2) Answer Decomposition<br>3) Categorizing Statements | Patient Questions | Comprehensiveness<br>Hallucination Rate |
| StaticKnowledge<br>[2] | 1) Knowledge Passage Question | Hospital Patient Clinics | Accuracy |
| Hallucinations-MIMIC-DI<br>[79] | 1) Hallucination Rate<br>2) Error Reduction<br>3) Quantitative Evaluation<br>4) Qualitative Evaluation | Doctor-written summaries<br>LLM-generated summaries | ROUGE F1 score<br>BERTscore<br>Relevance<br>Consistency<br>Fluency<br>Coherence<br>Simplification |
| MedHallBench<br>[245] | 1) Medical Visual QA<br>2) Image Report Generation | Textual case scenarios<br>Expert-annotated EMR<br>Radiology Images<br>Validation Exams | ACHMI indicators<br>(ACHMII & ACHMIS)<br>BertScore<br>METEOR<br>ROUGE<br>BLEU |

#### 4.1.1 Factual Verification

Factual verification techniques evaluate whether model-generated claims are supported by reliable evidence, either by assessing factual accuracy at the level of fine-grained facts [38] or at a more granular, claim-specific level [132]. The sub-component decomposed from the complex claims [38] provides fine-grained evaluation, while “atomic facts” supports granular evaluation, which is not an evaluation on the entire sentences [132]. By isolating individual or gross facts, this method ensures rigorous verification of multi-faceted medical claims.

#### 4.1.2 Summary Consistency Verification

Given the central role of summarization in clinical practice, ranging from condensing complex patient histories in electronic health records to integrating findings from clinical studies, it is essential to ensure that generated summaries accurately and faithfully represent the original information. Methods that evaluate summary consistency play a key role in detecting medical hallucinations, which occur when important clinical details are omitted, distorted, or fabricated during the summarization process.

Summary consistency methods evaluate whether a generated summary faithfully reflects the source content, categorized into **question-answering (QA)-based** and **entailment-based** approaches. QA-based methods assess consistency by generating questions from either the source or the summary. Recall-based method evaluates the summary based on question generated from the text [166], while consistency-based method [205] detects hallucination within summary by generating question from the summary and comparing factual inconsistencies. Both recall and consistency based detection could be used as a combination [163] and generate questions from both the source and the summary. Entailment-based methods use natural language inference to determine whether each sentence in the summary is logically entailed by the source. Another approach reranks candidate summaries based on entailment scores, detecting subtle inconsistencies that may not be captured by QA-based methods [61]. This approach emphasizes logical coherence and is well-suited for identifying hallucinations where facts are misrepresented or distorted. QA-based methods focus on fact recall, while entailment-based methods emphasize logical consistency, providing complementary approaches for hallucination detection.

#### 4.1.3 Uncertainty-Based Hallucination Detection

Uncertainty-based hallucination detection assumes that hallucinations occur when a model lacks confidence in its outputs. These methods rely on either **sequence log-probability** or **semantic entropy** to quantify uncertainty. Sequence probability-based methods detect hallucinations by analyzing the probability assigned to generated sequences. For example, [69] computes the log-probability of the sequence and flags low-probability outputs as potential hallucinations. Later work refines this approach by focusing on token-level probabilities and their contextual dependencies, improving hallucination detection by adjusting for overconfidence in certain predictions [248]. In contrast, semantic entropy-based methods shift the focus to the variability in meaning across different outputs. These approaches sample multiple LLM generations per query and cluster candidate generations into semantic equivalence groups; then, they assign a higher likelihood of hallucination to outputs with high semantic entropy [59]. More recent methods propose to alternatively probe how stable the LLM’s answer is under paraphrased versions of the same query, helping separate uncertainty caused by unclear question phrasings from uncertainty due to the model’s own knowledge gaps [76]. For medical LLMs, this helps determine whether the model requires further training on specific clinical concepts or if users need to be more precise in formulating their queries. Together, sequence probability and semantic entropy methods offer complementary approaches for hallucination detection, with sequence log-probabilities providing a general token-level uncertainty measure and semantic entropy capturing how stable the underlying meaning is.

### 4.2 Methods for Evaluating Medical Hallucinations

To effectively evaluate and quantify hallucinations in medical LLMs, we propose a systematic framework that aligns with the taxonomy presented in Table 2. This framework encompasses multiple measurement approaches, each addressing specific aspects of hallucination detection and evaluation across different healthcare applications. Specific tests designed to detect these hallucinations in clinical contexts are presented in Table 4.

**Table 4:** Benchmark tests for detecting hallucinations in LLM-generated clinical reasoning. This table outlines various evaluation tests designed to assess the reliability of LLMs in clinical contexts. Each test targets a specific challenge, such as maintaining chronological orders, summarizing complex cases, disambiguating medical jargon, identifying contradictory evidence, interpreting lab results, and generating differential diagnoses. In Section 7, we conduct chronological ordering test, 2) lab test understanding and 3) Differential Diagnosis Generation Test on LLM responses annotated by human physicians.

| Test Name | Objective | Description/Prompt |
| --- | --- | --- |
| **Chronological Ordering Test** | Detects if the LLM can maintain the correct sequence of events, which is crucial in medical diagnosis and treatment timelines. | You are a medical assistant who analyzes the medical record and organizes the information in temporal order. Given the presentation of the case, please organize the key medical events, including symptoms, test results, diagnoses, and treatment, with the corresponding times or durations (e.g., weeks of gestation, specific dates). Return the result in a dictionary format. |
| **Summarization Test** | Summarize this clinical case, including a chronological summary, diagnosis, and treatment plan for the patient. | Evaluates the LLM’s ability to condense and represent critical information without missing essential details or introducing extraneous information. |
| **Medical Jargon Disambiguation Test** | Explain any ambiguous or potentially confusing medical terms from this case report. | Tests if the LLM can accurately explain complex medical terms without creating confusion or offering incorrect definitions. |
| **Contradictory Evidence Test (CET)** | Is there any evidence or information in this case report that suggests multiple possible diagnoses? | Assesses whether the LLM can recognize conflicting information that might lead to different interpretations, thus testing its understanding of differential diagnoses. |
| **Lab Test Understanding: Detecting Abnormal Values** | Summarize the lab tests performed in this patient case. | You are a medical assistant who analyzes the Laboratory Data for the below medical case. [CASE GIVEN]. Given this, please provide your insights to the Laboratory Data |
| **Lab Test Understanding: Correlating Lab Values with Symptoms** | How do these lab results relate to the patient’s condition? | You are a medical assistant who analyzes the medical record with the laboratory data. [CASE GIVEN] Given this, please provide your insights to the Laboratory Data and correlate with the symptoms of this patient. |
| **Differential Diagnosis Generation Test** | Which cases could be considered when diagnosing a patient? | Based on the differential diagnosis of this patient, draw a knowledge-graph type of tree using json format following the differential diagnosis process in the case. |

**Table 5:**
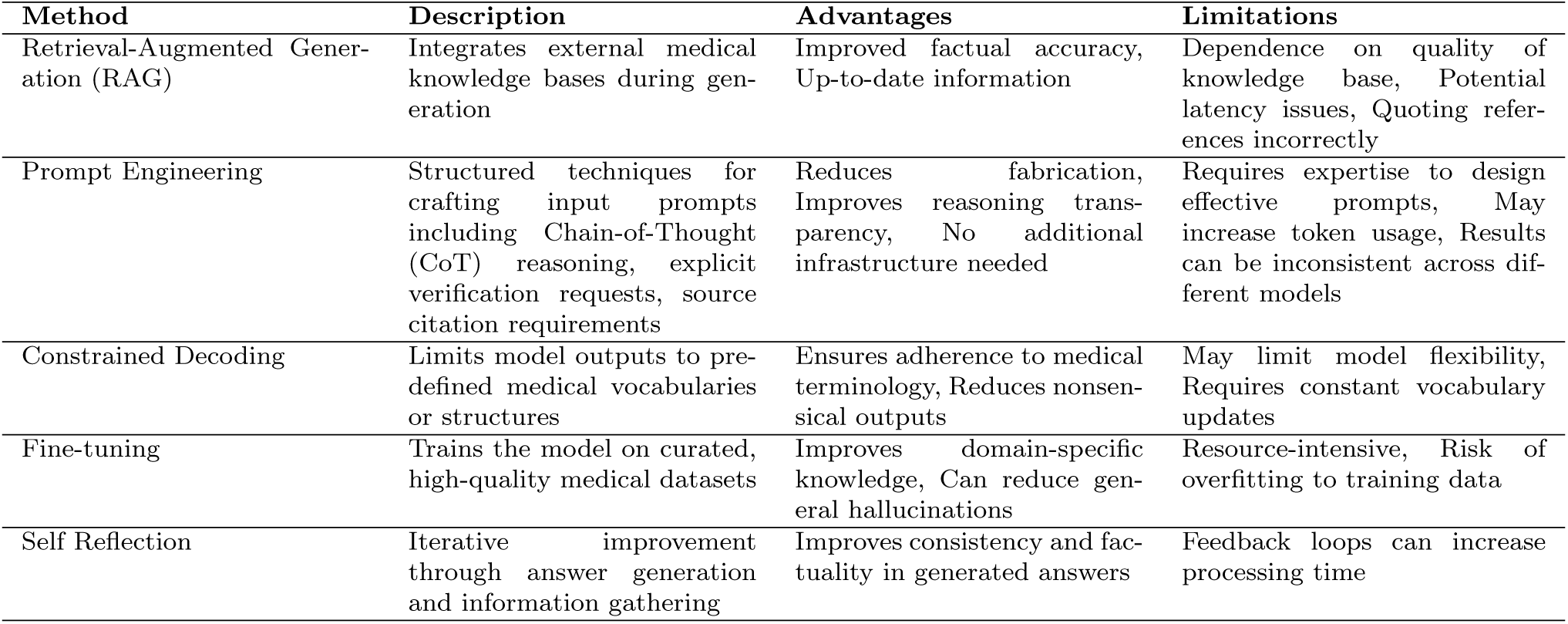
Strategies for mitigating medical hallucinations in LLMs. Methods include RAG, prompt engineering, constrained decoding, fine-tuning, and self-reflection, each addressing different aspects of factual accuracy, reasoning transparency, and domain specificity.

#### Factual Accuracy Assessment

This fundamental measurement approach directly addresses Factual Hallucinations and Research Hallucinations by comparing LLM outputs against authoritative medical sources. When an LLM generates information that contradicts established medical knowledge, it indicates the model is fabricating content rather than retrieving and applying accurate information. This involves both automated metrics (entity overlap, relation overlap) and expert verification, with particular emphasis on medical entity recognition and relationship validation. For instance, in Drug Discovery applications, this would assess whether drug-protein interactions described by the LLM align with established biochemical knowledge [95].

#### Consistency Analysis

This approach employs Natural Language Inference (NLI) and Question-Answer Consistency techniques to detect Decision-Making Hallucinations and Diagnostic Hallucinations in Clinical Decision Support Systems (CDSS) and EHR Management. Internal contradictions in an LLM’s response suggest the model is generating information without maintaining a coherent understanding of the medical case, indicating hallucination rather than reasoned analysis [178]. Future benchmarks and metrics are recommended to examine logical consistency across medical reasoning chains and evaluates whether treatment recommendations align with provided patient information and clinical guidelines.

#### Contextual Relevance Evaluation

This measurement method addresses Context Deviation Issues using n-gram overlap and semantic similarity metrics to assess whether LLM outputs maintain appropriate clinical context. When an LLM’s response deviates significantly from the medical query or context provided, it suggests the model is generating content based on spurious associations rather than addressing the specific medical situation at hand. This is especially important in Patient Engagement and Medical Education & Training applications, where responses must align precisely with the specific medical scenario or educational objective under consideration [4, 213].

#### Uncertainty Quantification

This approach uses sequence log-probability and semantic entropy measures to identify potential areas of Clinical Data Fabrication and Procedure Description Errors. High uncertainty in the model’s outputs, as indicated by low sequence probabilities or high semantic entropy, suggests the LLM is generating content without strong grounding in its training data, making hallucination more likely. This is particularly crucial in Medical Research Support and Clinical Documentation Automation, where fabricated information can have serious consequences [7, 203].

#### Cross-modal Verification

This measurement technique specifically targets Multimodal Integration Errors by evaluating whether textual descriptions are actually supported by the medical images or data they claim to describe [44]. When an LLM generates text about medical images that describes features or findings not present in the actual image, this indicates the model is hallucinating content rather than performing accurate interpretation [178]. The measurement process involves generating multiple descriptions of the same medical image, then using an evaluator model to compute entailment scores between the generated description and a ground truth imaging report. This approach is particularly critical for applications like Medical Question Answering and Clinical Documentation Automation where LLMs must generate accurate descriptions of medical imaging or laboratory results.

Each measurement approach may require both automated metrics and expert validation, with specific adaptations for medical domain requirements. For example, entity overlap metrics can be enhanced to specifically measure text similarity in medical terminology, procedures, and relationships, while NLI classifiers can be fine-tuned on medical literature and clinical guidelines.

### 4.3 Challenges in Medical Hallucination Detection

One of the fundamental challenges in detecting hallucinations lies in the ambiguity of the term itself, which encompasses diverse errors and lacks a universally accepted definition [83]. This makes it difficult to standardize benchmarks or evaluate detection methods effectively. Once a consensus on the definitions of hallucinations is established, there remains the issue of evaluating these phenomena effectively. Developing principled metrics aligned with a clear taxonomy of hallucinations is essential for advancing detection approaches. Often,domain-specific solutions tailored to the complex reasoning processes in medical contexts, such as entailment framework specific to radiology report, is useful to evaluate whether generated statements align with input findings [178].

#### Lack of a Reliable Ground Truth

A significant obstacle in hallucination detection is the frequent absence of, or the high cost of collecting a reliable ground truth, especially for complex or novel queries. For example, when simulating diagnostic tasks in radiology or summarizing complex patient histories, it is challenging to define what constitutes a *hallucination* without pre-established labeled examples [80]. This gap hinders both the evaluation of detection methods and the supervised training of models for hallucination detection of medical LLMs.

Annotating diagnosis recommendations for real-world medical cases is challenging in part due to Hickam’s Dictum [17], which recognizes that patients can have multiple coexisting conditions. Symptoms often align with various potential diagnoses, making it difficult to confidently determine a comprehensive diagnosis without dedicating significant time to the case. Given time constraints, medical annotation for evaluating AI hallucinations typically limits the time doctors have to assess each AI-generated output. This restriction makes it difficult to distinguish between clear AI hallucinations, or errors, and potentially useful, yet unconventional, diagnoses that may warrant further investigation.

Lastly, some clinical cases exhibit significant disagreement among clinicians, making it difficult to condense them into a single annotation. This problem is exacerbated in highly specialized domains, such as oncology, evidenced by a recent study evaluating ChatGPT’s ability to provide cancer treatment recommendations against National Comprehensive Cancer Network (NCCN) guidelines [36]. In this work, full agreement among three oncologists occurred in only 61.9% of cases [36], highlighting the inherent complexity of assessing AI-generated medical outputs in specialized domains.

#### Semantic Equivalence and Its Role in Detection

Semantic equivalence is the alignment of meaning between two pieces of text, ensuring they convey the same intended message [58, 147]. In the context of LLMs, this is used to identify inconsistencies or hallucinations by comparing multiple outputs sampled from the same input for contradictions or self-inconsistencies [59]. Additionally, semantic equivalence can verify whether a model-generated medical report accurately reflects a reference report [178]. In many cases, this is done using bidirectional entailment, a mutual verification process where each text is evaluated to confirm it logically supports, and is supported by, the other [59, 147]. However, there is a lack of comprehensive testing to determine whether entailment approaches perform effectively in specialized healthcare domains.

## 5 Mitigation Strategies

### 5.1 Data-Centric Approaches

As the capabilities of LLMs continue to evolve, there is a growing emphasis on data-centric approaches to improve their performance and reduce hallucinations. The quality, scope, and diversity of training data are fundamental to building reliable and accurate models, particularly in specialized fields such as biomedicine. This section outlines key strategies for reducing hallucinations through specialized dataset development and data augmentation techniques.

#### 5.1.1 Improving Data Quality and Curation

Pretrained LLMs, such as GPT-3 [19], GPT-4 [142], PaLM [33], LLaMA [183], and BERT [48], have demonstrated remarkable advancements, largely due to the extensive datasets used in their training. However, achieving high accuracy in biomedical applications requires fine-tuning on domain-specific corpora that are curated for quality, diversity, and task specificity. Models such as Flan-PaLM [33, 171] show high performance in medical benchmarks, illustrating the potential of targeted pre-training in improving LLM capabilities for complex medical reasoning and text generation. Fine-tuned LLaMA family models for medical tasks also show outstanding capability in the question-answering domain [212] and cross-language adaptability [210]. This progress emphasizes how domain-specific training can significantly improve LLM capabilities in handling advanced medical tasks.

Enhancing data quality and curation is critical for reducing hallucinations, as inaccuracies or inconsistencies in training data can propagate errors in model outputs. To address this, curated datasets have been developed to meet the specific requirements of specialized medical tasks. For example, MEDITRON-70B [29] is designed to improve medical reasoning, while MedCPT [89] enhances biomedical information retrieval, both of which are based on curated datasets for their respective purposes. These domain-specific datasets enable LLMs to develop a deeper understanding of medical knowledge, ultimately increasing reliability and accuracy. By leveraging high-quality, well-annotated datasets, models can better align with real-world clinical decision-making and minimize hallucinations that arise from incomplete or misleading information.

#### 5.1.2 Augmenting Training Data

Augmenting training data has become important in enhancing the reasoning capabilities of LLMs in medical applications. Augmentation techniques help bridge knowledge gaps, improve generalization, and mitigate biases in LLM-generated outputs. Several LLM-driven solutions have been introduced to enrich training datasets with clinically relevant information. For example, models used in patient-trial matching [238] improve compatibility between electronic health records (EHRs) and clinical trial descriptions, thereby refining model accuracy in real-world clinical settings. Furthermore, models like DALL-M [77] use a multistep process to generate clinically relevant characteristics by synthesizing data from medical images and text reports, allowing for more personalized healthcare solutions. Another prominent model, GatorTronGPT [153], trained on a comprehensive set of clinical data, improves the generation of biomedical text, facilitating the augmentation of medical training data for various tasks and downstream training applications.

### 5.2 Model-Centric Approaches

Model-centric approaches focus on directly improving LLMs through advanced training techniques and post-training modifications. Unlike data-centric methods that enhance the input data, these approaches aim to refine the model’s internal representations, reasoning capabilities, and output generation processes. Such methods are crucial in the medical domain, where factual accuracy, reliability, and interpretability are paramount for safe and effective clinical decision-making.

#### 5.2.1 Advanced Training Methods

##### Preference Learning for Factuality

To better align model outputs and behaviors with human preferences, several methods have been introduced, including direct preference optimization [DPO; 159], reinforcement learning from human feedback [RLHF; 145], and AI feedback [RLAIF; 114], utilizing techniques like proximal policy optimization [PPO; 175] as a training mechanism.

In addition to aligning outputs with human preferences, these methods have been extended to improve factuality by incorporating knowledge-based feedback signals [170, 184]. For instance, reinforcement learning from knowledge feedback [RLKF; 233] specifically trains models to generate accurate responses or reject questions when outside their knowledge scope, achieving superior factuality compared to decoding strategies or supervised fine-tuning [184].

Despite their potential, preference tuning demands a substantial volume of high-quality preference labels [114], posing a significant challenge for adoption in the medical field due to the high cost of annotations, limited number of expert annotators, and privacy concerns [228]. To address this, the use of synthetic data generated by LLMs with clinical knowledge has been proposed. For example, Mishra et al. demonstrate that using synthetic factual edit data from LLMs can effectively guide factual preference learning, resulting in more accurate outputs without the need for extensive human annotations.

#### 5.2.2 Post Training Methods

##### Model Knowledge Editing

Knowledge editing techniques provide a targeted approach to refining LLM outputs without requiring complete retraining. Unlike continual learning, which updates models through iterative fine-tuning, knowledge editing directly modifies model weights or adds new knowledge parameters [251]. A common approach is to train a model editor, which identifies and applies corrections to internal model representations to produce factually accurate outputs [47, 123]. This method offers efficiency by avoiding costly retraining, but it has significant drawbacks: it can inadvertently degrade model performance on unrelated tasks, struggles with applying multiple simultaneous edits, and often fails with edits that require broader contextual understanding [133].

Due to the complexity of medical knowledge and the lack of domain-specific benchmarks, knowledge editing has seen limited adoption in healthcare. Alternatively, parameter-efficient approaches add new modules, such as layer-wise adapters [231], instead of altering the base model directly. Such methods are more modular and less disruptive to overall model behavior. This promising framework also propose a domain-specific benchmark, the Medical Counter Fact (MedCF) dataset, to evaluate model edits in medical contexts, to evaluate model edits in medical contexts, demonstrating the effectiveness of targeted model editing in improving factual accuracy without compromising generalization.

##### Critic Models

In addition to training an LLM to produce more factual outputs, an auxiliary critic model can be used to critique the model’s outputs to re-prompt or edit its generation [122, 150]. Some works use self-refining methods, using the model itself to both critique and refine its own output [51, 97, 137], with the aim of improving the robustness of LLM reasoning processes to reduce hallucination. Although showing some promising results, these methods rely on prompting at each intermediate reasoning step and LLM’s reasoning capabilities to correct itself, which can result in unreliable performance gains [73, 120].

### 5.3 External Knowledge Integration Techniques

External knowledge integration techniques enhance the capabilities of LLMs by incorporating up-to-date and specialized information from external sources. These approaches are particularly valuable in the medical domain, where timely, accurate, and evidence-based information is crucial for reducing hallucinations and improving decision support.

#### 5.3.1 Retrieval-Augmented Generation

However, RAG itself can be a source of error if not implemented carefully. A primary failure mode is retrieval of irrelevant or low-quality information. For instance, if a query about a rare disease retrieves a forum post instead of a peer-reviewed article, the LLM may ground its answer in misinformation, leading to a confidently incorrect and potentially harmful response. Another failure mode is retrieval-generation conflict, where the retrieved document contradicts the model’s parametric knowledge, and the model struggles to reconcile the two, sometimes defaulting to its internal (and possibly outdated) knowledge.

Retrieval-augmented generation (RAG) [115] is a prominent method for integrating external knowledge without additional model retraining. The RAG process begins with the retrieval of relevant text and the integration of it into the generation pipeline [11], from concatenation to the original input to integration into intermediate Transformer layers [18, 84] and interpolation of token distributions of retrieved text and generated text [236].

In medical contexts, RAG has been shown to outperform model-only methods, such as CoT prompting, on complex medical reasoning tasks [224, 225]. More importantly, RAG’s ability to explicitly cite and ground outputs in retrieved knowledge makes it highly interpretable and controllable; qualities that are particularly valuable in clinical applications [154]. This has led to its adoption across various medical applications, including patient education [211], doctor education [243], and clinical decision support [214].

Specialized RAG frameworks tailored for healthcare further enhance the accuracy of LLMs by integrating medical-specific corpora and retrievers. For example, MedRAG, a systematic toolkit designed for medical question answering, combines multiple medical datasets with diverse retrieval techniques to improve LLM performance in clinical tasks [224]. Building on this, i-MedRAG (iterative RAG for medicine) introduces an iterative querying process where the model generates follow-up queries based on prior results. This iterative mechanism enables deeper exploration of complex medical topics, forming multi-step reasoning chains. Experiments show that i- MedRAG outperforms standard RAG approaches on complex questions from the United States Medical Licensing Examination (USMLE) and Massive Multitask Language Understanding (MMLU) datasets [225].

However, RAG techniques face key challenges that limit their effectiveness. The quality of generated responses heavily relies on the relevance and accuracy of retrieved documents. Poor retrieval results can propagate errors into model outputs [227]. Second, system maintenance overhead, i.e., curating and maintaining up-to-date retrieval corpora, especially for rapidly evolving fields such as medicine, requires significant resources [225]. Moreover, integrating misleading information from low-quality sources [109] or conflicting evidence [218] can degrade model performance and undermine trust in its outputs. Addressing these challenges requires advancements in retrieval models, knowledge base curation, and filtering mechanisms to ensure only high-quality, verified medical knowledge is incorporated into LLM outputs.

#### 5.3.2 Medical Knowledge Graphs

Knowledge graphs (KGs) have been used extensively to encode medical knowledge for LLMs and graph-based algorithms, especially in the medical domain [13, 34, 110, 235]. By structuring complex medical information into interconnected entities and relationships, KGs facilitate advanced reasoning and provide clear context and provenance [180], as each fact within the graph is traceable to its source [110] and informative through clear descriptions [34]. This traceability is particularly crucial in the medical field, where the accuracy and reliability of information are paramount.

The integration of KGs into LLMs has shown promise in mitigating hallucinations, instances where models generate plausible but incorrect information [110]. By grounding LLM outputs in the structured and verified data contained within KGs, the likelihood of generating erroneous or fabricated content is reduced in medical diagnosis. For instance, De Nicola et al. [54] highlight the potential of KGs to enhance diagnostic accuracy by encoding complex medical relationships and facilitating structured reasoning in clinical decision making. Similarly, Wang et al. [209] demonstrate how KGs can be applied to medical imaging, enabling the integration of multimodal data to reduce diagnostic errors in imaging analysis workflows. Yu et al. [239] explore how KGs support the management of chronic disease in children, providing actionable insights through data synthesis and predictive analytics. Furthermore, Gong et al. [70] focus on safe medicine recommendations, utilizing KG embeddings to mitigate risks associated with incorrect prescriptions. This synergy enhances the factual consistency of model outputs, a critical factor in medical applications where misinformation can have serious consequences. Recent studies have explored various methodologies to incorporate KGs into LLM workflows, aiming to improve the factual accuracy of generated content in tasks such as link prediction, rule learning, and downstream polypharmacy [65].

### 5.4 Uncertainty Quantification in Medical LLMs

A key dimension of reliability in large language models (LLMs) is their ability to detect and communicate when they are uncertain about a given query or piece of information. In clinical settings, where inaccurate or ungrounded outputs can mislead decision-making, robust mechanisms for uncertainty estimation are critical. When an LLM faces questions exceeding its familiarity or training scope, it ideally should communicate uncertainty or refrain from answering, rather than offering false confidence [111]. By quantifying how confident or uncertain a model is, LLMs can refrain from providing answers when knowledge gaps are significant [62, 86, 100, 215]. This section explores contemporary methods for modeling uncertainty, discusses their relevance to medical applications, and highlights the role of multi-LLM collaboration in reducing hallucinations.

#### 5.4.1 Methods for Confidence Estimation

Uncertainty in an LLM’s output can be represented and managed through various techniques, each addressing different dimensions of reliability.

**Model-Level and Training-Based Approaches.** Certain strategies integrate uncertainty estimation directly into the training process. For example, methods that introduce probabilistic layers or specialized loss functions can encourage models to produce calibrated confidence measures, rather than treating every prediction with equal certainty [86, 100]. Additionally, targeted knowledge integration during pretraining can reduce blind spots, although maintaining up-to-date domain coverage remains an ongoing challenge [62].

**Prompting and Post-Hoc Calibration.** Another class of methods focuses on refining uncertainty estimates after the initial model output. Prompt-based strategies encourage the LLM to self-assess its confidence, while post-hoc calibration techniques, such as temperature scaling or external calibrators, adjust logits or embedding representations [185, 215, 230]. Empirical findings indicate that such techniques can improve reliability in medical diagnosis tasks [67, 176], though larger models are not always better calibrated [49, 63, 168].

**Multi-LLM Collaboration.** Relying solely on a single LLM for self-correction can be risky if the model has learned skewed or incomplete patterns [91, 98, 229]. Multi-LLM collaboration attempts to reduce individual model biases by cross-verifying reasoning processes and outcomes. Voting or consensus-based approaches harness diverse model knowledge, mitigating hallucinations and overconfidence by highlighting discrepancies across peers [23, 52, 62, 240].

**Structured Uncertainty Sets.** In some instances, abstaining entirely may omit potentially valid insights, especially when a model has partial confidence. Techniques like conformal prediction strike a balance between outright abstention and blind certainty by providing sets of plausible answers with quantifiable error guarantees [1, 130]. Such methods let clinicians consider multiple options, each accompanied by a measure of confidence, rather than a single deterministic (and potentially incorrect) recommendation.

#### 5.4.2 Confidence Estimation in High-Stakes Applications

In high-stakes medical scenarios, reliably assessing uncertainty is paramount to preventing harmful outcomes. Models are often tasked with diagnosing critical conditions based on limited or ambiguous information. Direct prompts for confidence scores can help identify when the model is overstepping its reliable scope [185, 241], but further refinements are frequently required.

**Low-Resource Specialties as Illustrative Cases.** Some medical subfields, such as certain aspects of women’s health, suffer from limited training data and rapidly evolving guidelines [99]. While AI-driven tools can enhance patient education and self-care behaviors in these contexts, inaccuracies pose serious risks. Systems must therefore not only provide answers but also quantify and communicate any underlying uncertainties, improving the likelihood that clinicians and patients will seek validation for potentially flawed outputs [187, 221].

**Abstention and Deliberation.** When models generate multiple hypotheses without a single decisive answer, abstention thresholding allows them to refrain from giving conclusive guidance [64, 172] and instead guiding them to ask additional questions [111]. In some cases, multi-step or multi-agent deliberation can further refine uncertainty estimates, prompting the LLM to re-check facts or invite additional input [98, 229]. Such approaches reduce the risk of passing on guesswork as definitive advice, a particular concern in time-sensitive medical scenarios.

**Empirical Insights from Clinical Settings.** Recent work by Rodman et al. [155] demonstrated that GPT-4 can sometimes surpass clinicians in estimating disease likelihoods, yet both the model and human experts deviated substantially from actual prevalence rates. Meanwhile, exploration in embodied agents like Voyager [220] shows that advanced LLM frameworks often lack robust uncertainty quantification, even outside the medical domain. These findings underscore the broader relevance of uncertainty estimation: without systematic calibration, confident but unfounded responses can overshadow the potential benefits of AI in healthcare.

By designing models that effectively convey when they are uncertain; whether via post-hoc calibration, structured confidence sets, or consensus-driven deliberation; practitioners can better interpret and validate AI outputs. Such strategies are crucial for minimizing risk in clinical diagnostics, where the cost of error can be immediate and severe.

### 5.5 Prompt Engineering Strategies

Recent advances in medical LLM applications have demonstrated several prompting strategies for hallucination mitigation, each employing distinct cognitive frameworks to enhance diagnostic reliability. The **chain-of-medical- thought (CoMT)** [87] approach restructures medical report generation by decomposing radiological analysis into sequential clinical reasoning steps. Implemented for chest X-ray and CT scan interpretation, this method prompts models to first identify anatomical structures (*“Observe lung fields for opacities”*), then analyze pathological indicators (*“Assess bronchial wall thickening patterns”*), and finally synthesize diagnostic conclusions (*“Correlate findings with clinical history of chronic obstructive pulmonary disease”*). By mirroring radiologists’ diagnostic workflows through structured prompt templates, CoMT reduced catastrophic hallucinations by 38% compared to conventional report generation methods, as measured through the MediHall Score metric evaluating disease omission/fabrication rates.

The interactive **self-reflection methodology** introduces a recursive prompting architecture for medical question answering systems [97]. When deployed in clinical decision support scenarios, this approach initiates with a knowledge acquisition prompt (*“Generate relevant biomedical concepts for: {patient presentation}”*), followed by iterative fact-checking queries (*“Verify consistency between {generated concept} and current medical guidelines”*). For a case study involving rare disease diagnosis, this multi-turn prompting strategy improved answer entailment scores by 27% across five LLM architectures by forcing models to reconcile generated content with internal knowledge representations through prompts like *“Revise previous diagnosis considering [contradictory finding]”*. Human evaluations showed this reflection loop reduced critical hallucinations (misclassified disease types) by 41% in pediatric oncology use cases.

**Semantic prompt enrichment** [149] combines biomedical entity recognition with ontological grounding to constrain LLM outputs. Through integration of BioBERT for clinical concept extraction and ChEBI for chemical ontology alignment, this strategy appends verified domain knowledge directly to prompts. A representative implementation for drug interaction queries structures prompts as: *“Using ChEBI identifiers [CHEBI:48607=ibuprofen] and [CHEBI:35475=warfarin], describe metabolic pathway interactions considering CYP2C9 polymorphism risks”*. When applied to pharmacological report generation, this method reduced attribute hallucinations (incorrect dosage/formulation details) by 33% compared to baseline prompts, while maintaining 92% terminological consistency with FDA drug labeling databases. The technique demonstrates particular efficacy in oncology applications where precise molecular descriptor usage is critical - staging reports showed 29% fewer TNM classification errors when ontology-enriched prompts specified histological grading criteria.

**Chain-of-Knowledge (CoK)** [121] is a framework that dynamically incorporates domain knowledge from diverse sources to enhance the facutal correctness of LLMs. CoK generates an initial rationale while identifying relevant knowledge domains, and dynamically refines the rationale with knowledge from the identified domains to provide more factually correct responses. The authors evaluate CoK across various domain-specific datasets, including medical, physical, and biological fields, demonstrating an average improvement of 4.9% in accuracy compared to the Chain-of-Thought (CoT) baseline. To further validate the framework’s impact on reducing hallucinations, they present evidence of enhanced factual accuracy on single- and multi-step reasoning tasks using ProgramFC, a factual verification method based on Wikipedia. Additionally, human evaluations corroborate these findings, confirming that CoK consistently yields more accurate responses than the CoT baseline.

## 6 Experiments on Medical Hallucination Benchmark

### 6.1 Setup

While Section 5 outlined a broad range of mitigation strategies, this experimental evaluation section focuses on a representative subset; prompt engineering and retrieval-augmented methods chosen for their wide applicability and relevance in real-world clinical query systems.

We conducted a series of experiments to evaluate the effectiveness of various hallucination mitigation techniques on Large Language Models (LLMs) using the Med-HALT benchmark [152]. For Med-HALT, we sampled 50 examples from each of seven medical reasoning tasks (350 total cases). We compared the performance of several LLMs across different prompting strategies and retrieval-augmented methods. Our evaluation pipeline used UMLSBERTo **Medical Report:** AP and lateral views of the chest were provided in the X-ray. Lung volumes are low. There are findings consistent with chronic lung disease such as sarcoidosis. Prominence of the pulmonary interstitial markings is due to mild heart failure. There is no pleural effusion or pneumothorax. The size of the heart is normal. The cardiomediastinal silhouette is notable for a tortuous aorta. Bones are slightly osteopenic. The impression suggests that 1. Improving right upper lobe consolidation; 2. Mild heart failure; 3. Findings of chronic lung disease, most likely sarcoidosis. [138], a specialized medical text embedding model, to assess the semantic similarity between generated responses and the ground truth medical information. The following methods were implemented:

**Base:** This method served as our baseline. LLMs were directly queried with the questions from the Med-HALT benchmark without any additional context or instructions. This approach allows us to assess the inherent hallucination tendencies of the LLMs in a zero-shot setting. The prompt consisted solely of the medical question.

**System Prompt:** We prepended a system prompt to the user’s question. These system prompts were designed to guide the LLM towards providing accurate and reliable medical information, explicitly discouraging the generation of fabricated content. Examples of system prompts included instructions to act as a knowledgeable medical expert and to avoid making assumptions. However, it’s important to note that research [247] has questioned the consistent effectiveness of personas and system prompts in improving LLM performance on objective tasks. While our prompts aimed to enhance reliability, studies suggest that the impact of such prompts, particularly those relying on personas, might be variable and not always lead to significant performance gains.

**CoT:** We implemented Chain-of-Thought (CoT) prompting by appending the phrase “*Let’s think step by step.*” to each question. This encourages the LLM to articulate its reasoning process explicitly, which can improve accuracy by facilitating the identification and correction of errors during the generation process. This aligns with the principle of eliciting explicit reasoning steps from LLMs to enhance performance on complex tasks [219]. Furthermore, the generation of natural language explanations, as explored in work like e-SNLI [41], is related to CoT in its aim to make the model’s reasoning more transparent and potentially improve its factual correctness. This encourages the LLM to articulate its reasoning process explicitly, which can improve accuracy by facilitating the identification and correction of errors during the generation process.

**RAG:** We employed MedRAG [224], a retrieval-augmented generation model specifically designed for the medical domain. MedRAG utilizes a knowledge graph (KG) to enhance reasoning capabilities. For each Med-HALT question, we used MedRAG to retrieve relevant medical knowledge from the KG. This retrieved knowledge was then concatenated with the original question and provided as input to the LLM. This allows the LLM to generate responses grounded in external, validated medical information. We adapted publicly available MedRAG code and its associated KG for this implementation.

**Internet Search:** This approach leverages real-time internet search to provide LLMs with up-to-date information. We utilized the SerpAPIWrapper from langchain to perform Google searches.

### 6.2 Dataset & Tasks

We utilized the Medical Domain Hallucination Test (Med-HALT) benchmark [152] that has been specifically designed to assess and quantify hallucinations in LLMs within the medical domain. The Med-HALT benchmark employs a two-tiered approach, categorizing hallucination tests into:

- **Reasoning Hallucination Tests (RHTs):** These tests evaluate an LLM’s ability to reason accurately with medical information and generate logically sound and factually correct outputs without fabricating information. RHTs are further divided into:

– **False Confidence Test (FCT):** Assesses if a model can evaluate the validity of a randomly suggested “correct” answer to a medical question, requiring it to discern correctness and provide detailed justifications, highlighting its ability to avoid unwarranted certainty.
– **None of the Above (NOTA) Test:** Challenges models to identify when none of the provided multiple-choice options are correct, requiring them to recognize irrelevant or incorrect information and justify the “None of the Above” selection.
– **Fake Questions Test (FQT):** Examines a model’s ability to identify and appropriately handle non- sensical or artificially generated medical questions, testing its capacity to discern legitimate queries from fabricated ones.
- **Memory Hallucination Tests (MHTs):** These tests focus on evaluating an LLM’s ability to accurately recall and retrieve factual biomedical information from its training data. MHTs include tasks such as:

– **Abstract-to-Link Test:** Models are given a PubMed abstract and tasked with generating the corresponding PubMed URL.
– **PMID-to-Title Test:** Models are provided with a PubMed ID (PMID) and asked to generate the correct article title.
– **Title-to-Link Test:** Models are given a PubMed article title and prompted to provide the PubMed URL.
– **Link-to-Title Test:** Models are given a PubMed URL and asked to generate the corresponding article title.

By incorporating these diverse tasks, Med-HALT provides a comprehensive framework to evaluate the multifaceted nature of medical hallucinations in LLMs, assessing both reasoning and memory-related inaccuracies.

### 6.3 Metrics

#### Hallucination Pointwise Score

The *Pointwise Score* used in Med-HALT [152] is designed to provide an in-depth evaluation of model performance, considering both correct answers and incorrect ones with a penalty. It is calculated as the average score across the samples, where each correct prediction is awarded a positive score (*P_c_* = +1) and each incorrect prediction incurs a negative penalty (*P_w_*= *−*0.25). The formula for the Pointwise Score (*S*) is given by:

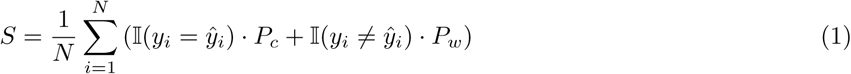

where:

- *S* is the final Pointwise Score.
- *N* is the total number of samples.
- *y_i_* is the true label for the *i*-th sample.
- *y*^*_i_* is the predicted label for the *i*-th sample.
- I(condition) is the indicator function, which returns 1 if the condition is true and 0 otherwise.
- *P_c_* = 1 is the points awarded for a correct prediction.
- *P_w_* = *−*0.25 is the points deducted for an incorrect prediction.

#### Similarity Score

The *Similarity Score* assesses the semantic similarity between the model’s generated responses and the ground truth correct answer, as well as the similarity between the response and the original question. This is achieved using UMLSBERT, a specialized medical text embedding model, and cosine similarity. The process is as follows:

1. **Embedding Generation with UMLSBERT:** For each question in the Med-HALT benchmark, and for each type of model output (Base, System Prompt, CoT, MedRAG, Internet Search), the following texts are encoded into embeddings using UMLSBERT:

1. The original medical *question*.
2. The *correct option* (ground truth answer).
3. The model’s generated *output* for each method.

2. **Cosine Similarity Calculation:** After obtaining the embeddings, the cosine similarity is calculated for each model output against two

references:

1. **Answer Similarity:** The cosine similarity between the embedding of the *correct option* and the embedding of the model’s *output*. This measures how semantically similar the generated response is to the ground truth answer.
2. **Question Similarity:** The cosine similarity between the embedding of the original *question* and the embedding of the model’s *output*. This evaluates how much the generated response is semantically related to the input question itself.
3. **Combined Similarity Score:** A *combined score* is then computed as the average of the *answer similarity* and the *question similarity* :

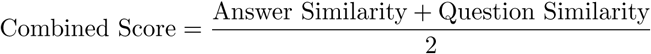

It is important to note that while the Pointwise and Similarity Scores provide quantitative measures of semantic correctness and alignment with ground truth, they do not directly capture the clinical safety or potential for patient harm. An output could be semantically similar but omit a critical warning, or factually correct on paper but clinically inappropriate. Therefore, these quantitative metrics are complemented by the qualitative analysis from physician annotators in Section 7, which specifically assesses the clinical risk associated with each hallucination.

### 6.4 Models

To assess medical hallucinations comprehensively, we evaluated a diverse set of foundation models, selected to represent a range of architectures, training paradigms, and domain specializations. Our selection included both general-purpose models, which represent the forefront of broadly applicable LLM technology, and medical-purpose models, designed or fine-tuned specifically for healthcare applications. This approach allowed us to compare hallucination tendencies across models with varying levels of domain expertise and general reasoning capabilities.

#### General-Purpose LLMs

These models are trained on large-scale datasets encompassing general text, code, and multimodal data, enabling broad applicability across diverse reasoning and generation tasks. Their inclusion establishes a baseline for hallucination performance and evaluates how general reasoning capabilities transfer to the medical domain. Within this category are models from OpenAI, Google, and DeepSeek. OpenAI models include o3-mini and o1, which allocate more inference time to deliberate reasoning before producing responses, improving performance on complex tasks such as scientific reasoning, coding, and mathematics. We also evaluate GPT-4o and GPT-4o mini, released in May and July 2024, respectively. GPT-4o is a multimodal model capable of processing and generating text, images, and audio with improved factual consistency, while GPT-4o mini offers a smaller, more efficient variant. The recently introduced GPT-5 extends this family, emphasizing advanced long-context reasoning and more reliable factual grounding. Google models include Gemini-2.5 Pro, designed for high-level multimodal reasoning and efficient cross-domain alignment between text and visual inputs. DeepSeek models feature DeepSeek-R1, a reasoning-optimized LLM that employs large-scale reinforcement learning on scientific and mathematical tasks to enhance logical consistency and reduce confabulation.

#### Medical-Purpose LLMs

These models are specifically adapted or trained for medical and biomedical tasks. Their inclusion is crucial for evaluating whether domain-specific training or fine-tuning effectively mitigates medical hallucinations compared to general-purpose models. This category includes PMC-LLaMA, Alpaca variants, and Google’s MedGemma. PMC-LLaMA is a model fine-tuned from LLaMA on PubMed Central (PMC), a free archive of biomedical and life sciences literature. It is designed to enhance performance in medical question answering and knowledge retrieval by leveraging a large corpus of peer-reviewed medical research papers. Alpaca Variants include AlphaCare-13B, an Alpaca-style LLaMA-based model further fine-tuned on medical question-answering datasets to improve clinical reasoning and dialogue capabilities, and MedAlpaca-13B, fine-tuned on a combination of medical datasets including clinical text and QA corpora, optimized for diverse medical NLP tasks. MedGemma is a specialized open-source multimodal model built on the Gemma 3 architecture, which incorporates research and technology derived from Google’s proprietary Gemini models. It is fine-tuned on biomedical literature, clinical text, and paired medical image and text data to support multimodal medical reasoning and structured report generation, emphasizing factual grounding and interpretability. This makes it particularly suitable for evaluating hallucination behavior and reliability in medical contexts.

### 6.5 Results

#### Reasoning Models Maintain Better Hallucination Resistance

Our experimental results, visualized in Figure 5, reveal that advanced reasoning models continue to excel in hallucination prevention, with the performance gap widening substantially in the latest generation. Notably, gemini-2.5-pro achieves hallucination resistance (97.9% with CoT, 87.6% baseline), while o3-mini (80.4% baseline) and deepseek-r1 (86.6% baseline) demonstrate robust performance that substantially exceeds o1 (64.0% baseline) and earlier-generation models like gpt-4o (54.4% baseline) and gpt-4o-mini (48.3% baseline).

The sustained superiority of general-purpose models suggests a fundamental architectural insight; hallucination resistance in specialized medical contexts emerges not from domain-specific fine-tuning, but from the sophisticated reasoning capabilities, internal consistency mechanisms, and broad world knowledge developed during large-scale pretraining. This finding has profound implications for medical AI development strategies, suggesting that investments in general intelligence capabilities may yield greater safety dividends than domain-specific optimization alone. The performance trajectory from gpt-4o-mini to gemini-2.5-pro (a 81.4% relative improvement) demonstrates that hallucination mitigation is a tractable problem that scales with model sophistication, offering optimism for achieving human-level reliability in medical AI systems.

#### CoT Demonstrates Robust Effectiveness with Statistical Evidence

Examining the impact of prompting strategies with enhanced statistical rigor, Chain-of-Thought (CoT) reasoning emerges as the most consistently effective intervention for mitigating hallucinations. CoT demonstrated significant improvements in 71% of models tested (*p <* 0.05), with 64% retaining significance after Benjamini-Hochberg FDR correction (*q <* 0.05). Crucially, System Prompting provides complementary gains, often working synergistically with CoT to further reduce hallucination rates exemplified by o3-mini (baseline 80.4% *→* System Prompt 81.4% *→* CoT 90.7%) and deepseek-r1 (baseline 86.6% *→* System Prompt 84.5% *→* CoT 90.7%).

To address multiple comparison concerns across 46 within-model tests, we applied Benjamini-Hochberg FDR correction. After correction at *q <* 0.05, 19 of 22 originally significant comparisons remained significant (86.4% retention rate), with all 11 highly significant results (*p <* 0.001) retaining significance. This high retention rate provides strong evidence that CoT effects represent genuine improvements rather than statistical artifacts. Only 3 borderline results (0.02 *< p <* 0.04) lost significance: o3-mini + system prompt (*p* = 0.024, *q* = 0.055), alpacare + system prompt (*p* = 0.030, *q* = 0.065), and gemini-2.5-pro + CoT (*p* = 0.034, *q* = 0.072).

The mechanistic basis for CoT’s effectiveness likely involves explicit reasoning traces that (i) surface intermediate steps for self-verification, (ii) reduce reliance on spurious pattern matching, and (iii) enable models to recognize and correct potential factual errors before committing to final outputs. The finding that even state-of-the-art models benefit from CoT suggests that reasoning transparency remains valuable even as base capabilities improve, with implications for clinical deployment where interpretability and error detection are paramount.

#### Semantic Similarity Stratifies Model Performance and Reveals Conceptual Understanding as a Key Mechanism

Analysis of similarity scores in Figure 5-b reveals striking performance stratification across model architectures. The highest-performing models: gemini-2.5-pro, o3-mini, and deepseek-r1 cluster in the high similarity range (0.8–0.9), indicating strong semantic alignment with ground truth medical information. Advanced reasoning model gpt-5 similarly positions in this elite tier (71.2% baseline resistance, similarity *>* 0.8). Conversely, medical-specific models (pmc-llama, medalpaca, alpacare, medgemma) consistently exhibit low similarity scores (0.1–0.5) alongside substantially higher hallucination rates.

This visual separation between medical-purpose models (clustered at low similarity and resistance) and general-purpose models (spanning higher similarity and resistance) reveals a critical insight: hallucination resistance correlates more strongly with *depth of conceptual understanding* as measured by semantic similarity to ground truth than with exposure to domain-specific training data. The failure of medical-specialized models to achieve high similarity scores despite explicit medical pretraining suggests they may be memorizing surface-level medical terminology without developing the deeper relational understanding necessary for reliable reasoning about complex clinical scenarios.

This finding challenges the prevailing assumption that domain specialization inherently improves medical AI reliability. Instead, our results suggest that models achieving genuine conceptual understanding (reflected in high similarity scores) naturally produce more factually accurate outputs, regardless of whether that understanding derives from general pretraining or medical fine-tuning. The implication is profound: effective medical AI may require not narrow domain optimization, but rather the sophisticated reasoning and knowledge integration capabilities that emerge from large-scale general intelligence development.

#### Domain-Specific Training Limitations Persist Despite Medical Pretraining

The results provide robust statistical evidence for a persistent and clinically meaningful performance gap between general-purpose and medical-specialized models. Despite explicit training on medical corpora, domain-specific models: pmc-llama (40.8% baseline), medalpaca (32.0% baseline), alpacare (28.6% baseline), and medgemma (52.6% baseline) achieve near or less than half the hallucination resistance of general-purpose models. Quantitatively, general-purpose models demonstrated substantially higher median baseline resistance than medical-purpose models (76.6% vs. 51.3%, average difference 25.2%, 95% CI: [18.7%, 31.3%], *U* = 27.0, *p* = 0.012, two-tailed Mann–Whitney test, rank-biserial *r* = *−*0.64, 95% CI: [*−*0.86, *−*0.28]).

The convergence of large effect size, adequate power, narrow confidence interval excluding zero, and independent ordinal evidence indicates a substantial and clinically meaningful difference that transcends statistical borderline status. From a clinical AI deployment perspective, this finding suggests that current medical-specialized models may not yet achieve the reliability standards necessary for unsupervised clinical use, whereas advanced general-purpose models are approaching acceptable thresholds.

We propose that the general-purpose model advantage stems from superior abstraction capabilities developed during diverse pretraining. Medical-specialized models, trained predominantly on medical literature, may overfit to domain-specific surface patterns without developing the flexible reasoning required to navigate novel clinical scenarios or detect logical inconsistencies. General-purpose models, exposed to broader reasoning contexts during pretraining, appear better equipped to apply first-principles reasoning and cross-domain knowledge integration— capabilities crucial for avoiding hallucinations in complex medical reasoning tasks. This suggests that the path to reliable medical AI may paradoxically require *less* domain specialization and *more* investment in general reasoning infrastructure.

***Search-Augmented Generation Reveals Differential Benefits Across Model Sophistication Tiers*** The effectiveness of search-augmented generation (SAG) demonstrates systematic variation across model capability levels, revealing important insights about when external knowledge retrieval provides value. At the highest capability tier, gpt-5 achieves the performance of 87.6% with search augmentation, representing a substantial +16.5% improvement over its baseline. This hallucination resistance suggests that even the most advanced models benefit from real-time access to authoritative external information, particularly for rapidly-evolving medical knowledge where internal parametric knowledge may be outdated.

Intriguingly, medical-specialized models show the largest *relative* gains from search augmentation despite maintaining lower *absolute* performance. For instance, pmc-llama exhibits a +46.1% relative improvement (40.8% baseline *→* 59.9% with Search, *p* = 1.9 *×* 10*^−^*^12^, *q* = 8.8 *×* 10*^−^*^11^ after FDR correction), suggesting that external retrieval can partially compensate for limited internal knowledge. However, even with search augmentation, these models fail to match the baseline performance of advanced general-purpose models, indicating that retrieval alone cannot overcome fundamental reasoning limitations.

Conversely, some highly advanced models show minimal or even negative effects from search augmentation: deepseek-r1 (86.6% baseline *→* 84.3% with Search, *−*2.3%) and o1 (64.0% baseline *→* 69.4% with Search, +5.4%). This pattern suggests that sophisticated models with strong internal knowledge may be better served by relying on their parametric memory rather than integrating potentially noisy or contradictory external sources. The observation that search occasionally *degrades* performance in advanced models raises important questions about retrieval integration strategies: models may require enhanced mechanisms to assess source reliability and resolve conflicts between parametric and retrieved knowledge.

**Clinical implications:** These findings suggest a nuanced deployment strategy for medical AI systems. For lower-capability models or rapidly-evolving medical domains (e.g., emerging infectious diseases, cutting-edge treatments), search-augmented generation provides valuable performance boosts and helps maintain currency. However, for highly capable models in stable knowledge domains, forced retrieval may introduce unnecessary complexity and potential error sources. Future medical AI systems may benefit from adaptive retrieval strategies that dynamically determine when external knowledge access is likely to improve versus degrade response quality, potentially using model uncertainty estimates or knowledge freshness requirements as triggers for retrieval invocation.

## 7 Annotations of Medical Hallucination with Clinical Case Records

To rigorously evaluate the presence and nature of hallucinations in LLMs within the clinical domain, we employed a structured annotation process. We built upon established frameworks for hallucination and risk assessment, drawing specifically from the hallucination typology proposed by Hegselmann et al. [80] and the risk level framework from Asgari et al. [8] (Figure 6) and used the New England Journal of Medicine (NEJM) Case Reports for LLM inferences.

**Fig. 6:**
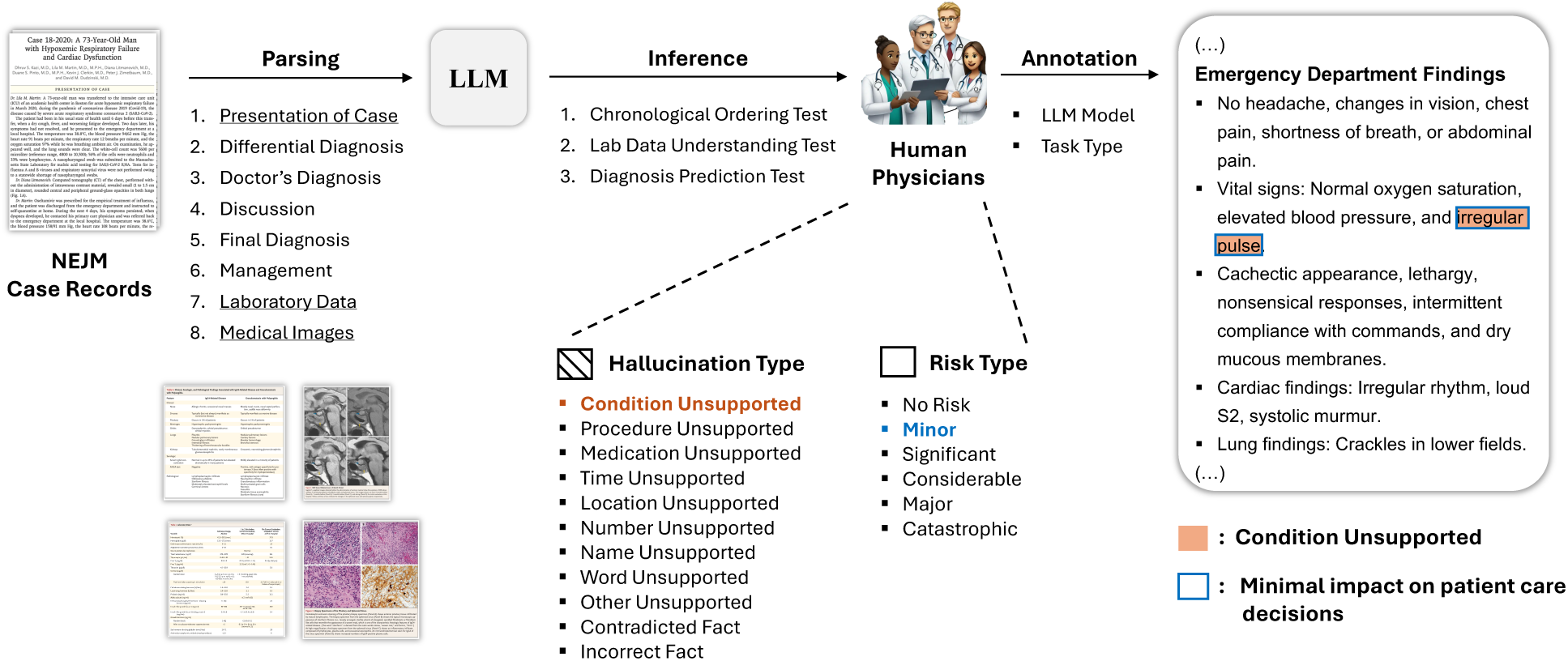
A**n annotation process of medical hallucinations in LLMs** (Section 7). We utilize New England Journal of Medicine (NEJM) case records, parsing them into key elements, and feeds them into the LLM for response generation. Physicians then annotate LLM-generated responses to identify medical hallucinations and potential risks, as exemplified by the inaccurate reporting of ‘irregular pulse’ in the patient’s Emergency Department findings.

**Fig. 7:**
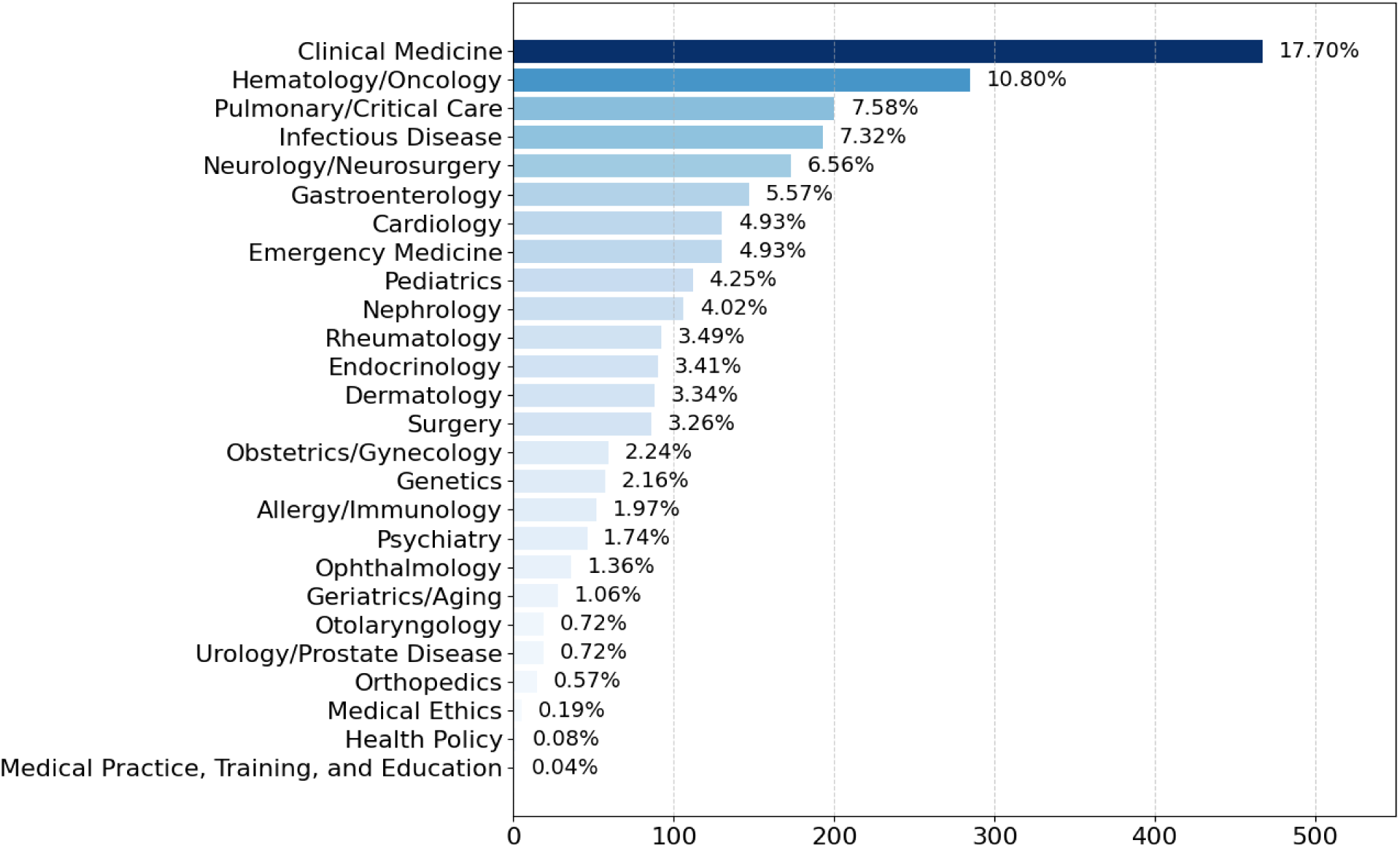
D**i**stribution **of medical specialties in sampled NEJM Case Records.** The figure illustrates the relative frequency of different medical domains, highlighting the predominant focus on clinical medicine, hematology/oncology, and pulmonary/critical care. The bar colors represent the proportion of cases within each specialty, revealing disparities in case representation across different fields.

### 7.1 Annotation Tasks for Detecting Medical Hallucination

We utilized the three representative evaluation tests outlined in Table 4 to probe LLMs for weaknesses in consistency, factual accuracy, and their ability to navigate the complexities and ambiguities inherent in clinical information (Figure 6). These tests were designed to elicit different types of potential hallucinations. When hallucinations occur, they can manifest in various forms, such as incorrect diagnoses, the use of confusing or inappropriate medical terminology, or the presentation of contradictory findings within a patient’s case.

Seven experienced annotators, each holding an MD degree or equivalent clinical expertise (including a geriatrician and an otolaryngologist), independently evaluated the generated outputs for each medical case record. Annotators were tasked with identifying and categorizing any hallucinations according to the types in Table 6 and assigning a corresponding risk level based on the definitions in Table 7. This rigorous annotation process allowed us to gain a granular understanding of the types and severity of hallucinations produced by LLMs in the medical domain.

**Table 6:** Categorization of medical hallucinations. This taxonomy, adapted from Hegselmann et al. [80], classifies different types of medical hallucinations based on their nature, including unsupported conditions, medications, procedures, temporal misrepresentations, and factual inconsistencies.

| Type | Description |
| --- | --- |
| Condition Unsupported | Mentions conditions not in context |
| Procedure Unsupported | References procedures not in context |
| Medication Unsupported | Mentions drugs/dosages not in context |
| Time Unsupported | Misrepresents temporal details |
| Location Unsupported | Fabricates/incorrect locations |
| Number Unsupported | Mismatched numerical values |
| Name Unsupported | Inaccurate/fabricated names |
| Word Unsupported | Generic/filler words not aligned |
| Other Unsupported | Miscellaneous errors, uncategorized |
| Contradicted Fact | Statements contradicting context |
| Incorrect Fact | Factual errors, not contradictions |

**Table 7:** Risk assessment framework for medical hallucinations. Adapted from Asgari et al. [7], this table categorizes potential risk levels of medical hallucinations, ranging from no risk (0) to catastrophic impact (5). The framework evaluates the severity of hallucinations based on their potential influence on clinical decision-making and patient safety.

| Level | Description |
| --- | --- |
| 0 | No Risk: No harm, no impact |
| 1 | Minor: Minimal impact on decisions |
| 2 | Significant: May affect decisions, unlikely harm |
| 3 | Considerable: Could lead to inappropriate decisions |
| 4 | Major: High probability of harmful decisions |
| 5 | Catastrophic: Could cause severe harm/death |

### 7.2 Dataset: The New England Journal of Medicine Case Reports

To evaluate the hallucination of LLMs, we used case records of the Massachusetts General Hospital, published in *The New England Journal of Medicine* (NEJM) [21]. We focused on the “Case Records of the Massachusetts General Hospital” series, filtering for Clinical Cases published between November 2000 and November 2018 to exclude cases related to the COVID-19 pandemic. This selection aimed to provide a representative sample of diverse medical conditions encountered in a major academic medical center. Leveraging NEJM’s categorization of medical specialties, we curated a dataset of 20 case reports, ensuring representation across a wide range of clinical areas. Each case was chosen to highlight certain challenges for the LLMs, e.g., complex differential diagnosis, detailed lab results. The distribution of cases across specialties, as defined by NEJM, is as follows (with the number of cases in the broader NEJM corpus for context):

Our smaller set of 20 cases sought to reflect this distribution, though not perfectly proportionally, to ensure coverage of common and less-frequent conditions.

### 7.3 Qualitative Evaluation of Clinical Reasoning Tasks Using NEJM Case Reports

To qualitatively assess the LLM’s clinical reasoning abilities, we designed three targeted tasks, each focusing on a crucial aspect of medical problem-solving: 1) chronological ordering of events, 2) lab data interpretation, and 3) differential diagnosis generation. These tasks were designed to mimic essential steps in clinical practice, from understanding the patient’s history to formulating a diagnosis.

#### Chronological Ordering Test

We first evaluated the LLM’s ability to sequence clinical events, a cornerstone of medical history taking and understanding disease trajectories. For this Chronological Ordering Test, we prompted the LLM with the question: *“Order the key events chronologically and identify temporal relationships between symptoms and interventions.”* Our results indicate that while the generated chronological ordering by the LLM was generally correct, it missed several important landmark events of the patient. For instance, in the case of opioid use disorder [217], crucial details regarding the patient’s history of oxycodone and cocaine use were absent from the generated timeline. These omissions are clinically significant as they provide context for the patient’s presentation. Furthermore, in the coronary artery dissection case [188], key treatment details, such as the administration of isosorbide mononitrate, were omitted. Precise temporal markers, such as specific dates for key events like hospital admission in the adenocarcinoma patient case [135], were also lacking. Lastly, we observed in [135] that the LLM struggled to group concurrent events, incorrectly treating them as separate, sequential occurrences.

#### Lab Test Understanding Test

Next, we stress-tested the LLM’s capacity to interpret laboratory test results and, critically, to explain their clinical significance in relation to the patient’s symptoms. For this Lab Data Understanding Test, we used the prompt: *“Analyze the laboratory findings and explain their clinical significance in relation to the patient’s symptoms.”* Our findings indicate that while the LLM could identify most laboratory results, it frequently failed to highlight and interpret abnormal values critical to understanding the patient’s conditions, particularly in cases like [135] and [105]. For example, in the adenocarcinoma patient case [135], the patient’s laboratory examination revealed a significantly elevated lactate level, a critical indicator of tissue hypoxia. Alarmingly, the LLM failed to report this crucial abnormality in its generated response, demonstrating a potential gap in its ability to prioritize and interpret clinically significant lab results.

#### Differential Diagnosis Test

Finally, we assessed the LLM’s ability to generate differential diagnoses, a vital skill in clinical decision-making. For the Differential Diagnosis Test, we asked: *“Based on the Presentation of Case, Lab Data and Images, what would be the possible diagnosis?”* When generating decision trees for differential diagnoses, the LLM was generally accurate in identifying primary considerations. However, it occasionally overlooked or minimized less obvious, yet clinically relevant, differential diagnoses. For instance, in the coronary artery dissection case [188], the LLM missed the potential for esophageal spasm as a differential diagnosis, highlighting a tendency to focus on the most prominent diagnoses and potentially overlooking a broader spectrum of possibilities.

### 7.4 Analysis of Hallucination Rates and Risk Distributions Across Tasks and Models

Based on expert annotations (Subsection 7.3), we rigorously quantified hallucination rates and the severity of associated clinical risks for five prominent LLMs: ‘o1’, ‘gemini-2.0-flash-exp’, ‘gpt-4o’,‘gemini-1.5-flash’, and ‘claude-3.5 sonnet’ (see Figure 1). Clinical risks were systematically categorized across a granular scale from ‘No Risk’ (0) to ‘Catastrophic’ (5), allowing for a nuanced evaluation of both the frequency and potential clinical ramifications of LLM-generated inaccuracies in medical contexts.

#### Overall Hallucination Rates and Task-Specific Trends

A notable task-specific trend emerged: Diagnosis Prediction consistently exhibited the lowest overall hallucination rates across all models, ranging from 0% to 22%. Conversely, tasks demanding precise factual recall and temporal integration – Chronological Ordering (0.25 - 24.6%) and Lab Data Understanding (0.25 - 18.7%) – presented significantly higher hallucination frequencies. This finding challenges a simplistic assumption that diagnostic tasks, often perceived as complex inferential problems, are inherently more error-prone for LLMs. Instead, our results suggest that current LLM architectures may possess a relative strength in pattern recognition and diagnostic inference within medical case reports, but struggle with the more fundamental tasks of accurately extracting and synthesizing detailed factual and temporal information directly from clinical text.

#### Model-Specific Hallucination Rates

GPT-4o consistently demonstrated the highest propensity for hallucinations in tasks requiring factual and temporal accuracy. Specifically, its hallucination rates in Chronological Ordering (24.6%) and Lab Data Understanding (18.7%) were markedly elevated compared to other models. Crucially, a substantial proportion of these hallucinations were independently classified by medical experts as posing ‘Significant’ or ‘Considerable’ clinical risk, highlighting not just the frequency but also the potential clinical impact of GPT-4o’s inaccuracies in these fundamental tasks. Interestingly, while GPT-4o’s hallucination rate in Diagnosis Prediction was also comparatively high in absolute terms (22.0%), it was marginally lower than that observed for Gemini-2.0-flash-exp (2.25%, although a potential data discrepancy between output logs and visual representation warrants further investigation).

The Gemini model family exhibited divergent performance characteristics. Gemini-2.0-flash-exp consistently maintained low hallucination rates across all three tasks, demonstrating relative strength in both factual/temporal processing and diagnostic inference. Its rates were notably low for Chronological Ordering (2.25%), Lab Data Understanding (2.25%), and Diagnosis Prediction (1.0%, with the aforementioned data discrepancy). In contrast, Gemini-1.5-flash displayed moderately elevated hallucination rates, particularly in Lab Data Understanding (12.3%) and Chronological Ordering (5.0%). While Gemini-1.5-flash also generated errors categorized as ‘Significant’ and ‘Considerable’ risk, these occurred at lower frequencies than observed with GPT-4o.

Claude-3.5 and o1 consistently emerged as the top-performing models across this evaluation, exhibiting the lowest hallucination rates across all tasks and risk categories. Remarkably, both models achieved a 0% hallucination rate in the Diagnosis Prediction task, suggesting a high degree of reliability for diagnostic inference within this specific context. Claude-3.5 demonstrated exceptionally low hallucination rates of 0.5% (Chronological Ordering) and 0.25% (Lab Data Understanding). o1 mirrored this strong performance, with equally low or slightly superior rates of 0.25% for both Chronological Ordering and Lab Data Understanding.

#### Risk Level Distribution and Clinical Implications

While GPT-4o’s higher hallucination frequency and associated clinical risk severity in Chronological Ordering and Lab Data Understanding tasks are concerning, the surprisingly low overall hallucination rates in Diagnosis Prediction; especially the 0% rate achieved by Claude-3.5 and o1 offer a nuanced perspective. These findings indicate that while current LLMs are not uniformly reliable across all clinical reasoning tasks, specific models, such as Claude-3.5 and o1, may possess a nascent but promising aptitude for diagnostic inference within structured medical case presentations. However, the consistent presence of ‘Significant’ and ‘Considerable’ risk errors even within models exhibiting lower aggregate hallucination rates underscores a critical and overarching implication: irrespective of overall performance metrics, the deployment of any current LLM in clinical settings necessitates rigorous, task-specific validation protocols, continuous performance monitoring, and careful integration within human-in-the-loop workflows. The potential for even low-frequency, but high-risk, hallucinations in fundamental tasks like temporal sequencing and factual recall necessitates a cautious and evidence-driven approach to LLM adoption in healthcare, prioritizing patient safety and clinical accuracy above claims of generalized AI proficiency.

### 7.5 Inter-rater Reliability Analysis

#### Quantifying Agreement in Hallucination Annotation

To rigorously assess the consistency and reliability of our qualitative evaluations, we conducted an inter-rater reliability analysis. Seven expert annotators, each holding an MD degree or advanced clinical specialization, independently evaluated the outputs generated by the language models for each of the 20 clinical case reports. The analysis focused on two key dimensions: (1) hallucination type and (2) clinical risk level. To quantify agreement among annotators, we employed the *Average Pairwise Jaccard-like Index* [85], a metric well suited for multi-label tasks where raters identify overlapping sets of items rather than mutually exclusive categories. This approach avoids the restrictive assumptions of Cohen’s *κ* and Fleiss’ *κ*, which require categorical exclusivity.

The aggregated inter-rater agreement, averaged across all cases and models, yielded a Jaccard-like Index of **0.272** for hallucination-type classification and **0.347** for clinical-risk-level assessment. These values correspond to a *moderate level of agreement* within the context of complex medical-text evaluation, consistent with previously reported ranges (0.25–0.40) for similarly nuanced clinical annotation tasks [104, 148]. In practical terms, this indicates that while annotators generally converged on the presence and severity of hallucinations, residual variability reflects the inherent interpretive difficulty of distinguishing clinically meaningful errors from stylistic or minor factual deviations.

#### Moderate Agreement in Medical Annotation

The observed Jaccard-like Index values (ranging from 0 to 1) thus represent a moderate degree of inter-rater agreement; not perfect consensus, but robust given the linguistic and clinical ambiguity of the task. This finding highlights the inherent challenge of achieving full uniformity when evaluating nuanced medical text for factual and clinical accuracy. As illustrated in Figure 8, annotators showed variability when assessing an LLM’s summary of a patient case: one expert identified the omission of a Roux-en-Y gastric bypass as a clear hallucination, while others interpreted discrepancies in the reported timelines of clinic visits differently. This example highlights how factual omissions with direct clinical impact are readily agreed upon, whereas subtler temporal or stylistic inconsistencies elicit subjective judgments, reflecting the layered nature of medical hallucination assessment.

**Fig. 8:**
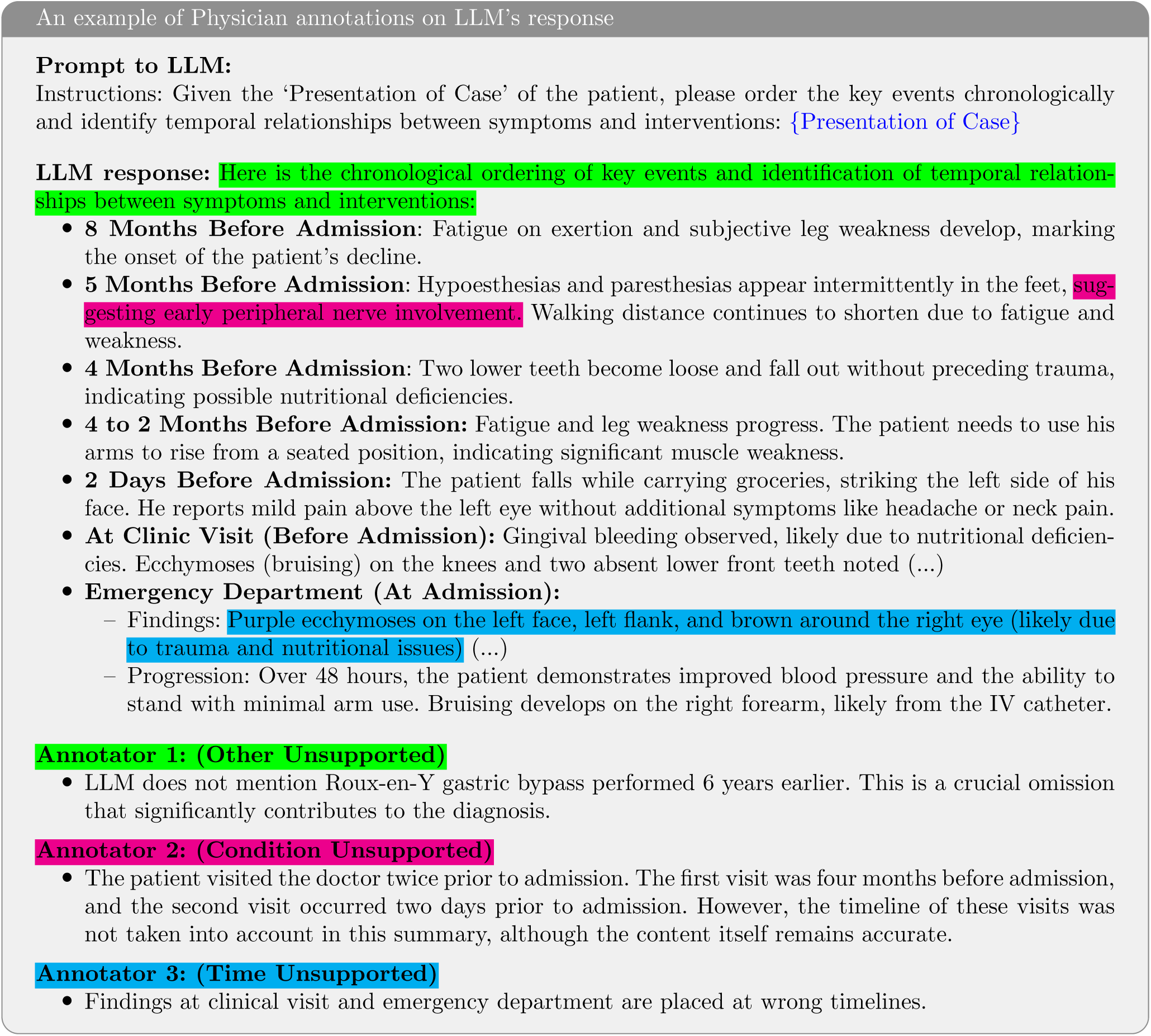
P**h**ysician **annotations on a GPT-4o generated chronological ordering of clinical events.** The original case can be found at https://www.nejm.org/doi/full/10.1056/NEJMcpc1802826.

#### Sources of Subjectivity and Annotation Challenges

Discussions with the expert annotators highlighted several key factors contributing to the observed inter-rater variability, many of which are exemplified in the annotation discrepancies shown in Figure 8. A primary challenge lies in the nuanced distinction between bona fide medical hallucinations and less critical errors, a point clearly illustrated by Annotator 1’s identification of the omitted Roux-en-Y gastric bypass. This omission represents a potentially clinically significant factual inaccuracy, arguably a ‘bona fide’ hallucination due to its relevance to patient history and diagnosis. In contrast, Annotators 2 and 3 focused on temporal inaccuracies, highlighting discrepancies in the timeline of doctor’s visits and the emergency department visit. These temporal issues, while potentially less clinically critical than the omission of a major surgery, still represent deviations from the source text and demonstrate the subjective interpretation of ‘accuracy’ in summarizing complex medical timelines. This distinction necessitates a high degree of clinical judgment, as annotators must decide not only if information is factually present but also its clinical relevance and the acceptable level of summarization detail.

#### Methodological Rigor to Enhance Annotation Consistency

To mitigate potential subjectivity and enhance annotation consistency, we implemented several methodological safeguards prior to and during the annotation process. We developed a comprehensive and interactive annotation web interface ^†^ (Figure B1, B2, B3) that incorporated detailed, operationally defined criteria and illustrative examples for each hallucination type and clinical risk level category. Moreover, we established proactive communication channels with each annotator, encouraging open dialogue and providing ongoing support to address any ambiguities, interpretational challenges, or uncertain cases encountered during their assessments. This iterative process aimed to refine understanding of the annotation guidelines and promote convergent interpretations among the expert raters.

#### Time Investment and Expertise Demands in Medical Annotation

The time investment required for annotation varied according to the annotator’s domain expertise and the specific complexity of each case report. Annotators were explicitly authorized and encouraged to leverage external, authoritative medical resources, including platforms such as UpToDate and PubMed, to inform their evaluations and ensure comprehensive assessments. The NEJM case reports, characterized by their depth, clinical intricacy, and frequent presentation of unusual or diagnostically challenging conditions, necessitated a substantial time commitment from each annotator for thorough and conscientious evaluation of the language model outputs.

#### Value of Qualitative Insights Despite Agreement Limitations

Despite the inherent challenges in achieving perfect inter-rater agreement in this complex domain, the patterns and trends emerging from our annotation process remain profoundly valuable. The observed inter-rater reliability, while moderate, is sufficient to support the identification of systematic biases and error modalities within the language models’ clinical reasoning and text generation capabilities. The qualitative insights derived from this rigorous annotation process offer critical directions for future model refinement and for developing strategies to mitigate clinically relevant medical hallucinations in large language models, as elaborated in the subsequent sections.

## 8 Survey on AI/LLM Adoption and Medical Hallucinations Among Healthcare Professionals and Researchers

To investigate the perceptions and experiences of healthcare professionals and researchers regarding the use of AI / LLM tools, particularly regarding medical hallucinations, we conducted a survey aimed at individuals in the medical, research, and analytical fields (Figure 9). A total of 75 professionals participated, primarily holding MD and/or PhD degrees, representing a diverse range of disciplines. The survey was conducted over a 94-day period, from September 15, 2024, to December 18, 2024, confirming the significant adoption of AI/LLM tools across these fields. Respondents indicated varied levels of trust in these tools, and notably, a substantial proportion reported encountering medical hallucinations—factually incorrect yet plausible outputs with medical relevance—in tasks critical to their work, such as literature reviews and clinical decision-making. Participants described employing verification strategies like cross-referencing and colleague consultation to manage these inaccuracies (see Appendix A for more details).

This study received an Institutional Review Board (IRB) exemption from MIT COUHES (Committee On the Use of Humans as Experimental Subjects) under exemption category 2 (Educational Testing, Surveys, Interviews, or Observation). The IRB determined that this research, involving surveys with professionals on their perceptions and experiences with AI/LLMs, posed minimal risk to participants and met the criteria for exemption.

The survey instrument, comprising 31 questions, was administered online, achieving a 93% completion rate from 75 participants. The resulting dataset of 70 complete responses formed the primary input for our analysis, enabling both quantitative and qualitative examination of the data. For example, qualitative analysis of open-ended responses on hallucination instances allowed us to identify recurring themes, which were then quantified to assess the prevalence and impact of medical hallucinations.

Respondents identified key factors contributing to medical hallucinations, including limitations in training data and model architectures. They emphasized the importance of enhancing accuracy, explainability, and workflow integration in future AI/LLM tools. Furthermore, ethical considerations, privacy, and user education were highlighted as essential for responsible implementation. Despite acknowledging these challenges, participants generally expressed optimism about the future potential of AI in their fields. The following subsections detail these findings, providing a comprehensive analysis of respondent demographics, tool usage patterns, perceptions of correctness and hallucinations, and perspectives on the future development and safe implementation of AI/LLMs in healthcare and research.development and safe implementation of AI/LLMs in healthcare and research.

### 8.1 Respondent Demographics

A total of 75 respondents participated in the survey, representing a diverse range of professional backgrounds within the medical and scientific communities. The largest group comprised Medical Researchers or Scientists (*n* = 29), followed by Physicians or Medical Doctors (*n* = 23), Data Scientists or Analysts (*n* = 15), and Biomedical Engineering professionals (*n* = 5), and others (*n* = 3). Most respondents held advanced degrees, with 52 possessing either a PhD or MD, 11 holding a Master’s degree, 9 with a Bachelor’s degree and 3 with others. Their professional experience varied: 30 had worked 1–5 years, 25 had 6–10 years, and 19 had over 20 years of professional experience. This breadth of expertise and educational attainment ensured that the survey captured perspectives across multiple career stages and levels of specialization.

### 8.2 Regional Representation

The geographic distribution of participants highlighted regions with robust AI infrastructures. Asia was most strongly represented (*n* = 27), followed by North America (*n* = 22), South America (*n* = 9), Europe (*n* = 8), and Africa (*n* = 4). While this sample provided valuable insights into regions where AI is more deeply integrated into healthcare and research workflows, the limited representation from other continents signals a need for future investigations to include a broader global perspective.

### 8.3 Usage and Trust in AI/LLM Tools

AI/LLM tools were well integrated into respondents’ routines: 40 used these tools daily, 9 used them several times per week, 13 used them few times a month, and 13 reported rare or no usage. Despite this widespread adoption, trust levels exhibited more caution. While 30 respondents expressed high trust in AI/LLM outputs, 25 reported moderate trust, and 12 indicated low trust. These findings suggest that although AI tools have gained significant traction, users remain mindful of their limitations, seeking greater reliability and interpretability.

### 8.4 Perceived Correctness and Encounters with AI Hallucinations

Perceptions of correctness were mixed. Of the 61 respondents, 21 believed that AI/LLM outputs were “often correct”, 18 stated they were “sometimes correct”, and 6 felt they were “rarely correct”. More critically, hallu- cinations—instances where the AI-generated plausible but incorrect information—were widely encountered by 37 respondents. Such hallucinations emerged across various tasks: literature reviews (38 mentions), data analysis (25), patient diagnostics (15), treatment recommendations (13), research paper drafting (16), grant writing (4), solving board exams (7), EHR summaries (4), patient communication (5), insurance billing (2), and citations (1). The prevalence of hallucinations in high-stakes tasks emphasized the need for meticulous verification and supplemental oversight.

### 8.5 Responses to AI Hallucinations

Respondents reported multiple strategies for addressing hallucinations. The most common approach was cross-referencing with external sources, employed by 85% (51) of respondents. Other strategies included consulting colleagues or experts (12), ignoring erroneous outputs (11), ceasing use of the AI/LLM (11), directly informing the model of its mistake (1), updating the prompt (1), relying on known correct answers (1), and examining underlying code (1). Assessing the impact of these hallucinations on a 1–5 scale, 21 respondents rated the impact as moderate (3), 22 rated it as low (2), 9 saw no impact (1), 5 observed high impact (4), and 2 reported a very high impact (5). This distribution suggests that while hallucinations are common, many respondents have developed coping mechanisms that mitigate their overall influence.

### 8.6 Causes of AI Hallucinations

Respondents identified various potential causes of hallucinations. Insufficient training data was the most frequently cited factor (31 mentions), followed by biased training data (31), limitations in model architecture (30), lack of real-world context (26), overconfidence in AI-generated responses (24), inadequate transparency of AI decision-making (14), and others (4). Moreover, 50 out of 59 participants believe the hallucination they experienced or observed might impact patient health. However, only 6 out of 59 participants believe there is more than a medium severity of the impact of AI hallucinations on their daily work, given AI has not yet fully merged into clinical workflow. These reflections underscore the multifaceted technical and ethical dimensions that must be addressed to improve model reliability.

### 8.7 Limitations of AI/LLMs and Future Outlook

lack of domain-specific knowledge emerged as the most critical limitation (30) followed by the privacy and data security concerns (25), accuracy issues (24), lack of standardization/validation of AI tools (23), difficulty in explaining AI decisions (21), a range of ethical considerations (20), and others (3).

Despite these concerns, the sentiment toward future developments was predominantly positive. A majority of respondents were optimistic (32) or very optimistic (24), with only a small minority expressing pessimism (3). Regarding the direct impact of AI/LLMs on patient health, opinions were mixed: 21 respondents believed there was an impact, 15 did not, 22 were uncertain, and 16 did not provide a clear stance. This divergence highlights an evolving field where the clinical value of LLMs is still being established.

### 8.8 Commonly Used AI/LLM Tools

In terms of specific technologies, ChatGPT was the most commonly mentioned tool (30 mentions), followed by Claude (20), Google Bard/Gemini (16), and open-source models such as Llama (15). Additional tools like Perplexity (9), Alphafold (2), Copilot (1), Scite and Consensus (1) also featured, reflecting a diverse ecosystem of AI solutions supporting medical and research professionals.

### 8.9 Future Priorities and Hallucination Safeguards

When asked about priorities for improvement, respondents emphasized enhancing accuracy (12 mentions), explainability (10), and ethical considerations, including bias reduction and privacy (8). Integration with existing tools (7) and improving speed and efficiency (3) were also noted. To safeguard against hallucinations, recommendations included manual cross-checking and verification (10), human supervision and expert review (8), confidence scoring or indicators (5), improving model architecture and training (5), training and education on AI limitations (4), and establishing ethical guidelines and standards (3). These suggestions outline a path toward greater reliability, transparency, and responsible integration of AI in healthcare.

## 9 Regulatory and Legal Considerations for AI Hallucinations in Healthcare

### 9.1 Deploying AI Systems in the Real-world Healthcare

AI systems developed to be deployed in the real-world need to be assessed for quality, safety, and reliability control [20] . Furthermore, they are required to meet codes of ethics and regulatory frameworks established by expert societies and governmental bodies, as models’ errors can lead to life-threatening consequences [30]. Although frameworks specifically designed for AI as a medical device are less developed compared to those for other types of medical devices, established rules (Figure 10 ) and new policies from expert associations and governmental organizations apply when an AI tool is used as a medical device. Incorporating these regulatory requirements during development aims to ensure that AI tools are effectively developed.

**Fig. 9:**
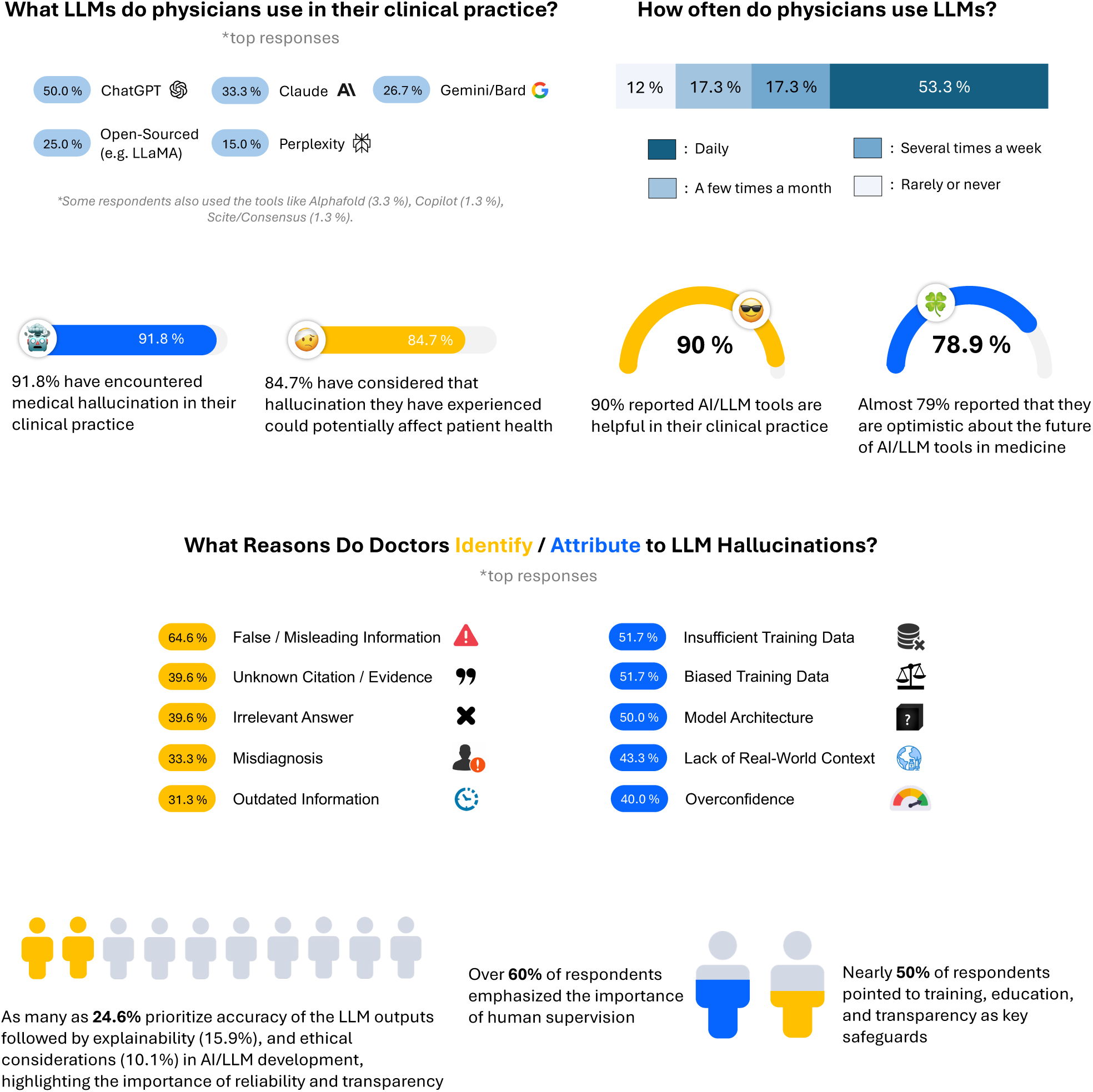
K**e**y **insights from a multi-national clinician survey on medical hallucinations in clinical practice.** The survey highlights the most commonly used LLMs and their frequency of use among physicians (top), clinicians’ experiences with LLM hallucinations and their perspectives on AI-assisted medical practice (middle), and the primary reasons attributed to LLM hallucinations along with the importance of human oversight, training, and transparency as safeguards (bottom).

**Fig. 10:**
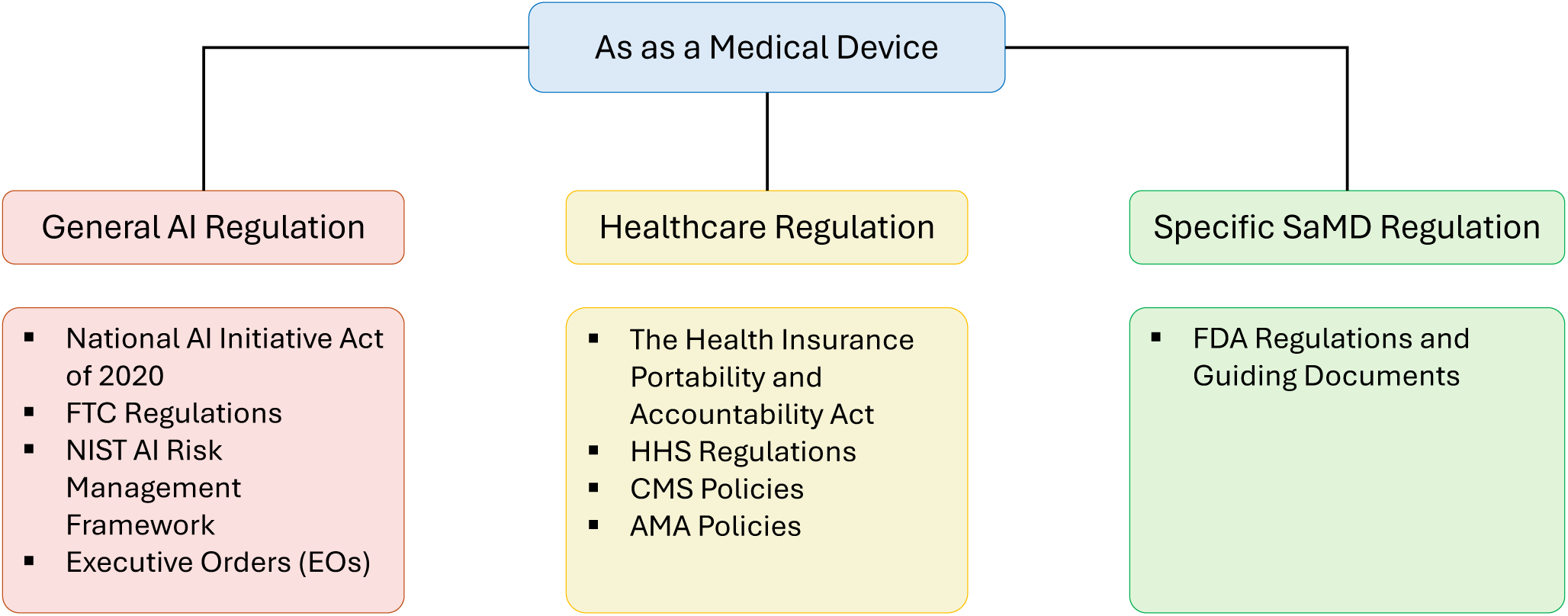
R**e**gulatory **Landscape for AI as a Medical Device:** A Framework Categorizing General AI, Healthcare, and Software as a Medical Device (SaMD)-Specific Regulations.

### 9.2 Federal Oversight of AI Systems

At the broadest level, the deployment of an AI system in the United States is governed by federal agencies tasked with ensuring public safety and consumer protection. The Federal Trade Commission (FTC) maintains primary oversight authority, with the power to take action against companies that misrepresent AI capabilities or deploy systems that cause consumer harm [57]. This oversight has become increasingly critical as the accessibility of AI development tools enables a wider range of entities to create and deploy AI systems, sometimes without adequate expertise or safety considerations. The regulatory landscape was significantly shaped by Executive Order 14110, signed in October 2023 [40], which established comprehensive requirements for AI system safety testing and transparency. This order works in conjunction with the Office of Science and Technology Policy’s Blueprint for an AI Bill of Rights [181] and the National Institute of Standards and Technology’s AI Risk Management Framework [140] to create a foundational structure for responsible AI development and deployment.

### 9.3 Healthcare-Specific Regulatory Requirements

When AI systems enter the healthcare domain, they encounter additional layers of regulation designed to protect patient safety and privacy. The Health Insurance Portability and Accountability Act (HIPAA) establishes fundamental requirements for the handling of protected health information [193]. Any AI system processing patient data must comply with HIPAA’s Privacy Rule [192], Security Rule [194], and Breach Notification Rule [191]. These requirements significantly impact how AI systems can access, process, and store medical data, with substantial penalties for non-compliance.

The American Medical Association (AMA) has also established specific guidelines for AI use in healthcare that emphasize transparency, physician oversight, and patient safety [10]. These guidelines explicitly position AI systems as augmentative tools rather than replacements for clinical judgment, maintaining that ultimate responsibility for medical decisions must remain with healthcare providers [9].

### 9.4 FDA Regulation of AI as Medical Devices

The Food and Drug Administration (FDA) plays a central role in regulating AI systems that function as medical devices. These systems are classified as Software as a Medical Device (SaMD) [199] when they are intended for use in the diagnosis, treatment, or prevention of disease. The FDA has established a risk-based framework for evaluating these systems, with three primary pathways for approval: (1) Premarket Approval (PMA) for high-risk devices [198]; (2) De Novo classification for novel moderate-risk devices; (3) 510(k) clearance for moderate to low-risk devices that are substantially equivalent to existing approved devices [195]

Moreover, recognizing the unique challenges posed by AI/ML-enabled medical devices, the FDA has published specific guidance documents, including Good Machine Learning Practice (GMLP) [196]. These guidelines address issues such as data quality, algorithm validation, and performance monitoring that are particularly relevant to AI systems.

### 9.5 State-Level Regulations

At the state level, new regulations are emerging to address AI-specific concerns. Colorado’s SB 24-205 [39] and California’s Assembly Bill 2013 [27] represent early efforts to regulate high-risk AI systems and ensure transparency in AI development.

### 9.6 Emerging Challenges in AI Healthcare Regulation

Although the regulatory framework for traditional medical AI applications is better established, generative AI systems present unprecedented challenges that strain existing oversight mechanisms. These systems’ unique charac- teristics—including their stochastic outputs, continuous learning capabilities, and complex integration with clinical workflows—create regulatory gaps that require innovative approaches [156].

Current frameworks, which designed for deterministic medical technologies, struggle to address several key aspects of generative AI systems. Unlike traditional medical devices with predictable outputs, generative AI systems can produce variable responses to identical inputs, making validation against ground truth particularly challenging. These systems often operate across both regulated and non-regulated applications, creating complex oversight scenarios [75]. Perhaps most critically, their ability to generate plausible but incorrect information poses unique safety risks, as demonstrated by our analysis of state-of-the-art systems (Figure 1) [30].

Regulatory bodies are beginning to adapt to these challenges. The FDA has introduced new approaches to change control for AI/ML-enabled medical devices [197], acknowledging the need for more flexible oversight of systems that continue to learn and evolve after deployment. However, these adaptations primarily address supervised learning systems rather than the unique challenges posed by generative AI. The development of effective regulatory frameworks for generative AI requires a data-driven approach that can quantify and categorize different types of hallucinations, establish clear risk thresholds for different clinical applications, and create protocols for monitoring and reporting AI-related adverse events.

### 9.7 Liability and Legal Framework

Finally, the integration of generative AI into healthcare also creates novel liability challenges that existing legal frameworks struggle to address. Traditional medical malpractice law, based on the concept of deviation from standard of care, faces significant hurdles when applied to AI-generated errors. When an AI system generates incorrect or misleading information, determining liability becomes complex, potentially involving AI developers and their training methodologies, healthcare providers using the system, and healthcare institutions implementing the technology [24].

The “black-box” nature of many AI systems further complicates the establishment of clear causal relationships between system outputs and patient harm [5]. This opacity challenges traditional legal requirements for establishing negligence and causation, particularly when multiple parties share responsibility in the technology’s lifecycle.

Legal scholars have proposed several frameworks to address these challenges. One approach involves expanding traditional malpractice standards to include specific requirements for AI system use, including mandatory critical evaluation of AI outputs and documentation of AI-assisted decision-making [5]. Another proposal suggests treating AI systems as products, with potential liability for systematic hallucinations or errors, though this approach faces challenges due to AI systems’ ability to evolve through continuous learning [117].

A particularly promising approach involves a distributed liability model that allocates responsibility based on stakeholder roles and control levels [56]. This framework emphasizes proportional responsibility distribution while encouraging comprehensive risk management protocols and structured validation procedures. Such an approach could incentivize all parties to maintain robust safety measures while promoting continued innovation.

The development of effective legal frameworks for AI in healthcare requires careful attention to informed consent, documentation standards, and causation criteria. Clear guidelines must be established for documenting AI-assisted decisions and determining responsibility in cases of adverse events. These legal considerations must evolve alongside technological advances to ensure that the benefits of AI in healthcare can be realized while maintaining robust patient safety protections.

## 10 Conclusion

In this study, we formally define and characterize *medical hallucination* as a reasoning-driven failure mode of foundation models, distinct from general hallucinations in both origin and clinical consequence. Through a comprehensive taxonomy and physician-audited benchmark, we reveal that most medical hallucinations stem not from missing medical knowledge, but from failures in causal and temporal reasoning. These errors, manifesting as mis- ordered symptom progression, flawed diagnostic logic, or misplaced causal inference persist even in large-scale models, indicating that greater parameter count and data coverage alone do not translate to safer clinical reasoning. Our empirical evaluation demonstrates that structured prompting and retrieval-augmented generation can reduce hallucinations by over 10%, yet high-risk reasoning errors remain, underscoring the limits of current architectures in approximating human clinical judgment. Complementing these findings, a global survey of clinicians revealed that over 90% had encountered AI-generated medical hallucinations, with the majority recognizing their potential to cause patient harm. Together, these results establish a mechanistic understanding of why hallucinations arise in medicine and how they differ fundamentally from errors in other domains. Moving forward, rigorous benchmarking against expert clinicians is essential to delineate which reasoning failures are uniquely algorithmic versus shared with human practitioners. Longitudinal evaluations in real-world clinical workflows will be critical to linking benchmark performance with patient outcomes. Ultimately, developing trustworthy medical foundation models will require integrating reasoning transparency, continuous evidence retrieval, and calibrated uncertainty estimation, ensuring that AI systems evolve from plausible responders to clinically accountable collaborators in patient care.

## Data Availability

Med-HALT is a publicly available dataset and NEJM Medical Records can be access after the sign-up.

https://www.nejm.org/browse/nejm-article-category/clinical-cases?date=past5Years

https://github.com/medhalt/medhalt

## Acknowledgements

The authors thank Chelsea Joe (MIT) for contributions to the initial brainstorming and the review of the paper on LLM hallucination and mitigation strategy. We also thank Rosalind Picard (MIT) for her insightful review and high-level comments that helped refine the paper. Additionally, we appreciate Yoon Kim (MIT) for his guidance on research direction and idea development. We also thank Peter Szolovits (MIT) for his pivotal role in conceptualizing medical hallucinations and contributing to the brainstorming process.

We extend our gratitude to Yanjun Gao (University of Colorado Anschutz Medical Campus) for their detailed review of Section 5, and to Monica Agrawal (Duke University) for her insightful feedback on the manuscript.

Furthermore, we would like to thank Leo Celi (MIT) for sharing his valuable research ideas, providing critical paper review, offering insightful perspectives on aspects of medical research, and contributing real-world practice stories and examples that enriched our understanding. We are grateful to Shannon Shen (MIT) for his helpful advice on paper review and guidance on research direction.

Finally, we are deeply indebted to Hyunsoo Lee, Kayoung Shim, and Yeji Lim from Seoul National University Hospital (SNUH) for their tireless efforts and dedication in annotating the NEJM Case Records. Their meticulous work required a significant investment of time and expertise, and we greatly appreciate their contributions.

This research was supported by a grant of the Korea Health Technology R&D Project through the Korea Health Industry Development Institute (KHIDI), funded by the Ministry of Health & Welfare, Republic of Korea (grant number : RS-2024-00439677)

## Appendix A Survey Details

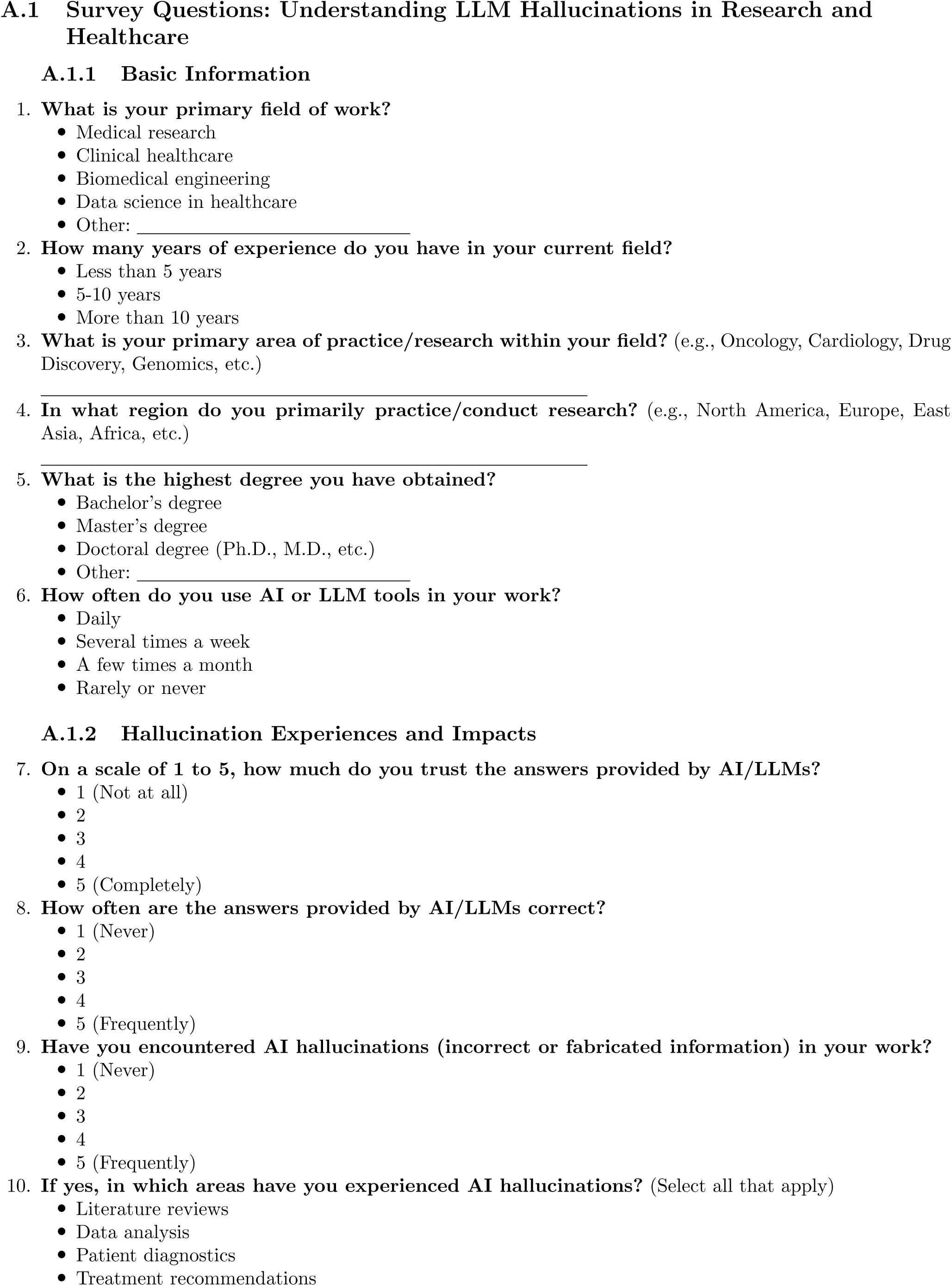
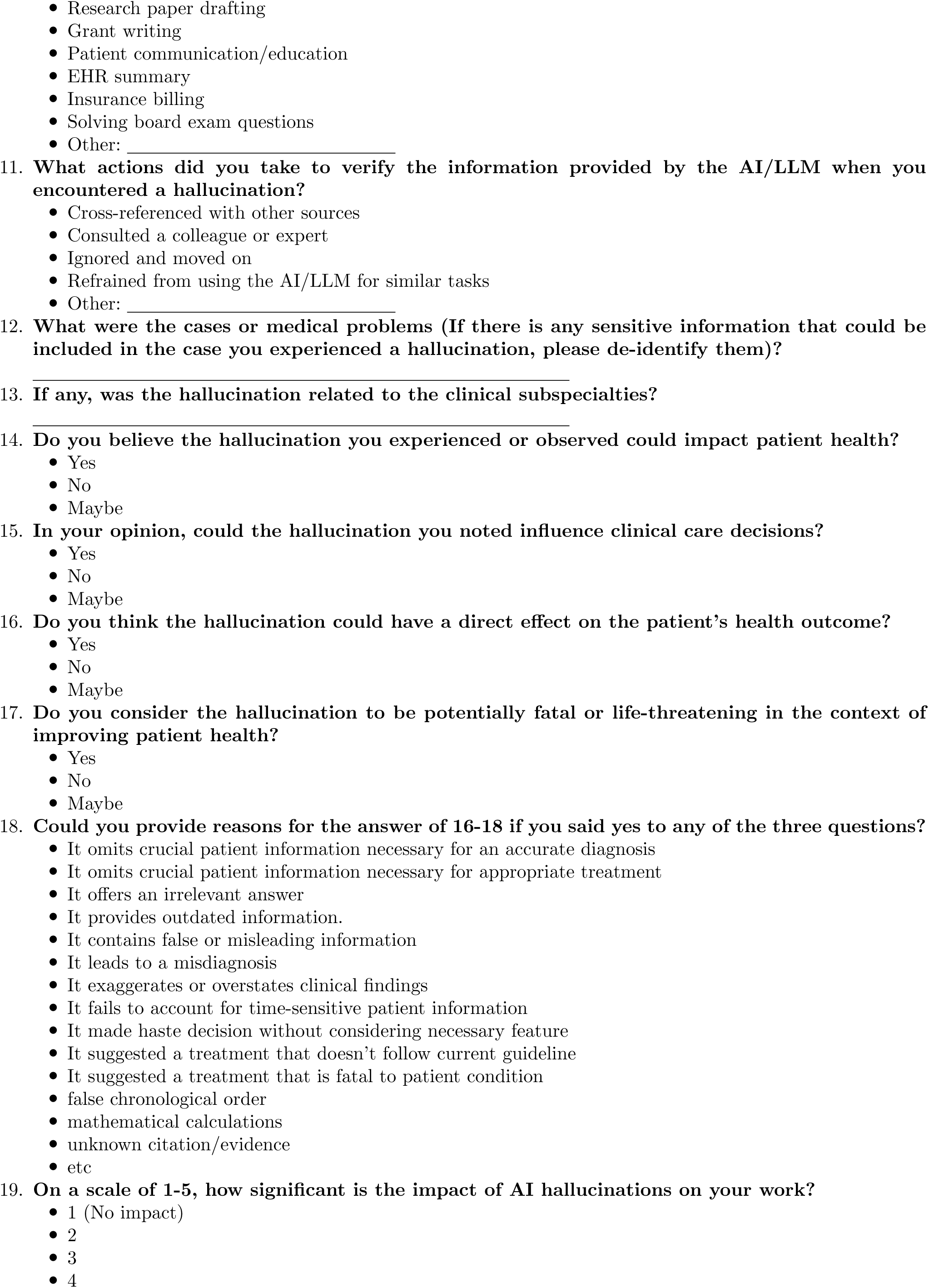
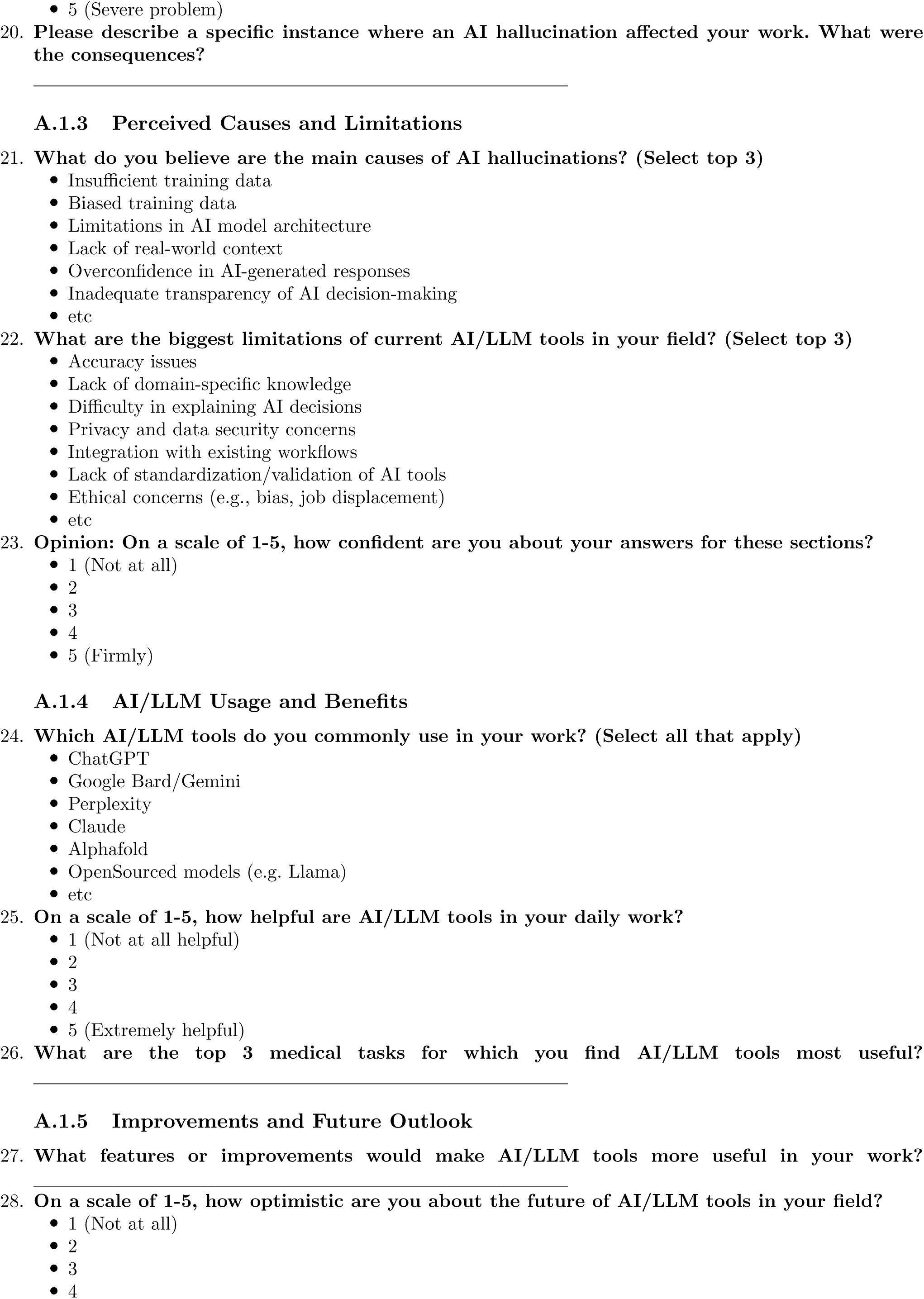
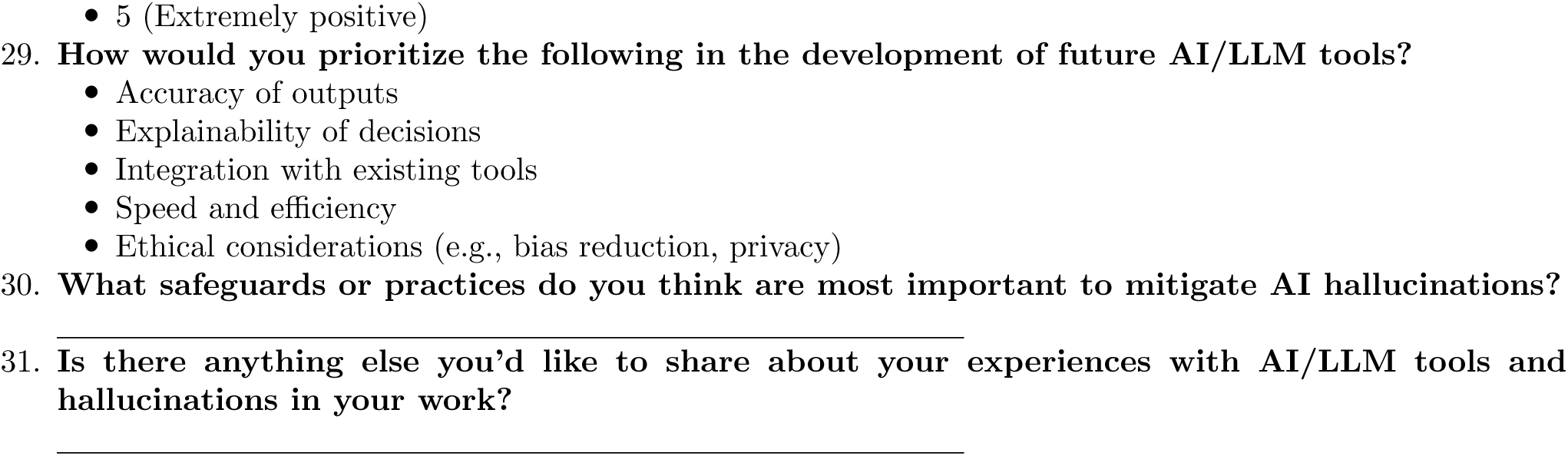
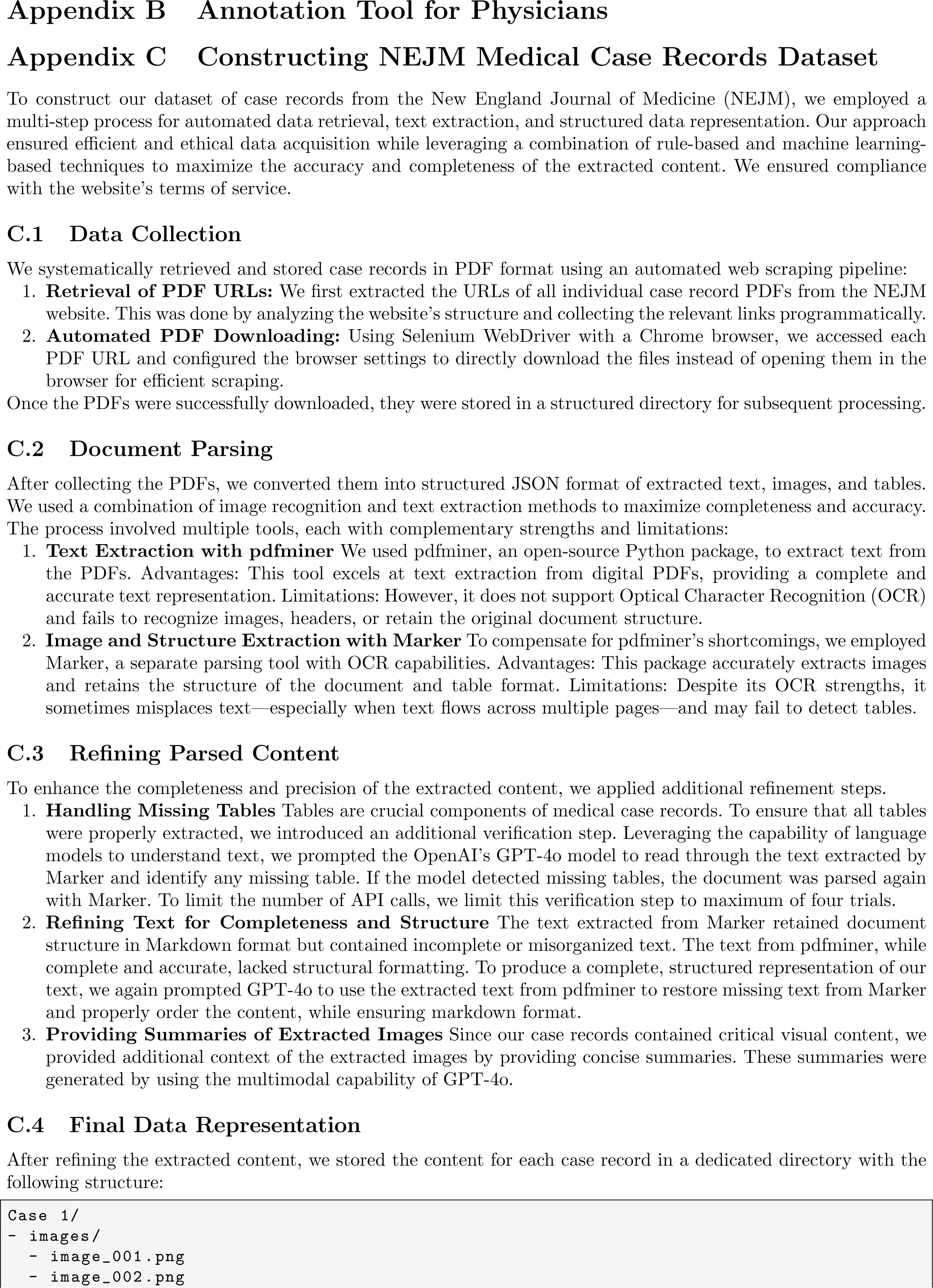
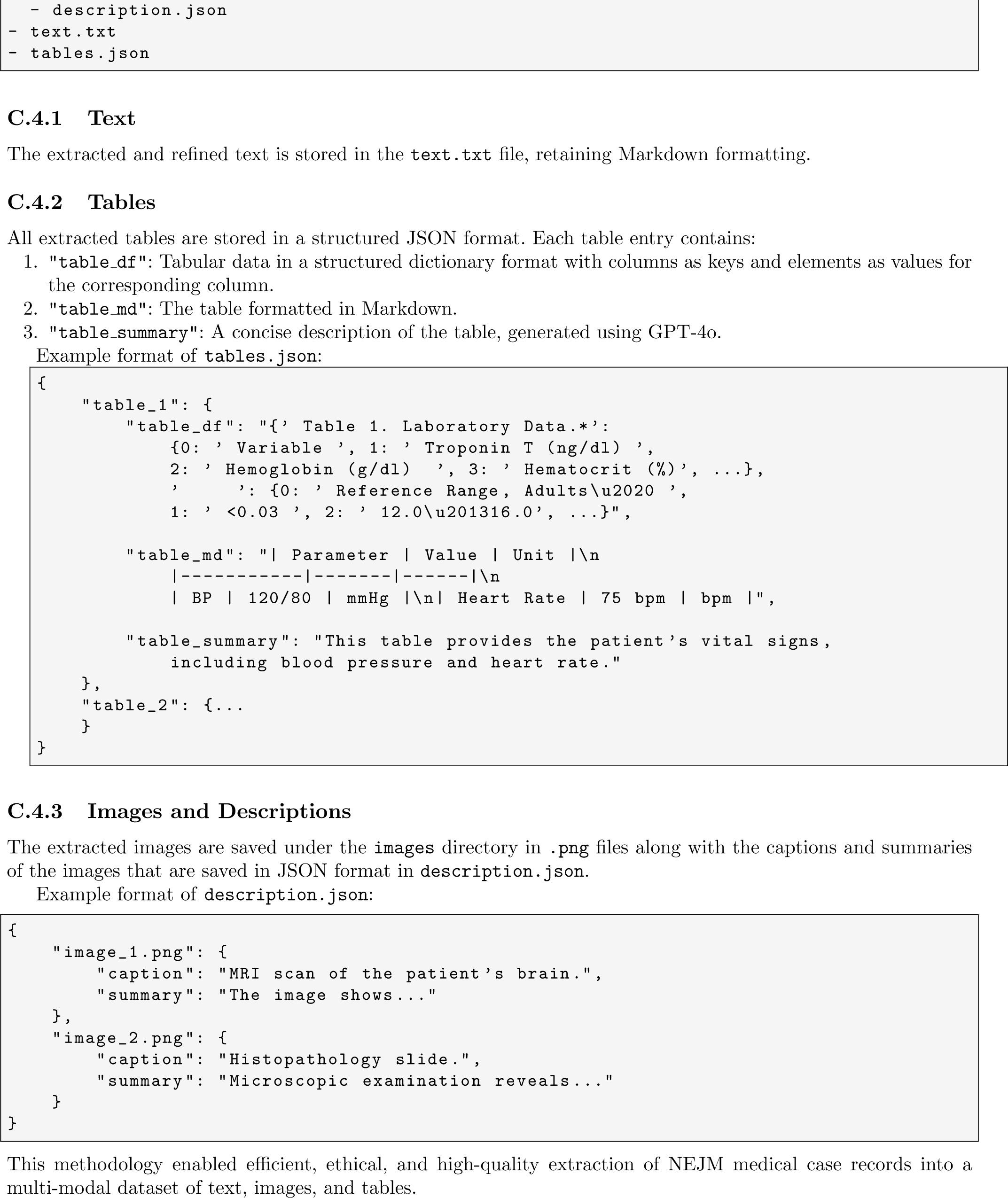

---

**Fig. B1:**
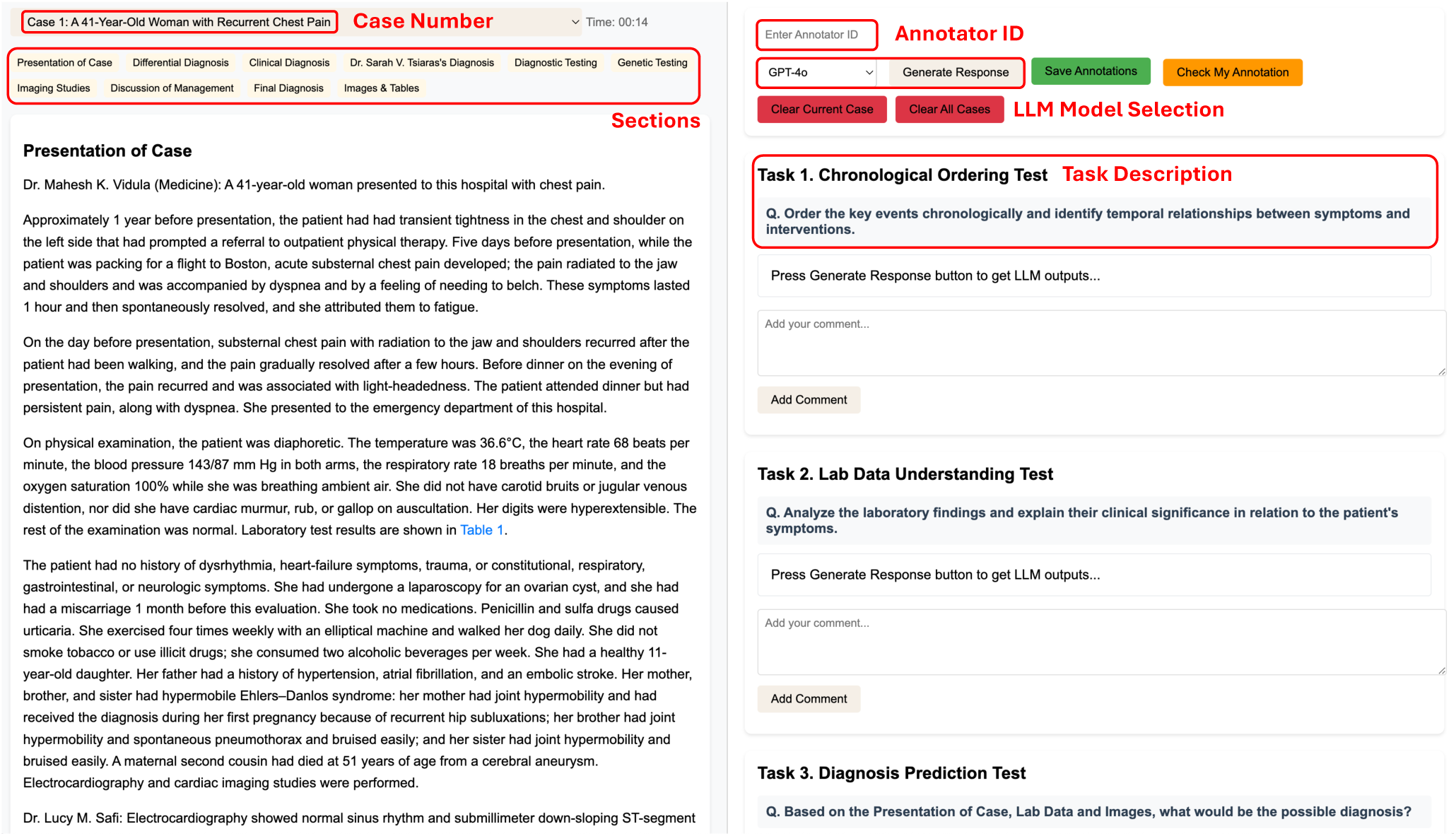
Web-based annotation tool for NEJM Case Records. The interface displays a clinical case with sections for case presentation, diagnosis, and testing. On the right, the tool provides annotation tasks for doctors, including chronological ordering of events, lab data understanding, and diagnosis prediction. Annotators can input their ID, select an LLM model, and save or check their annotations within the tool.

**Fig. B2:**
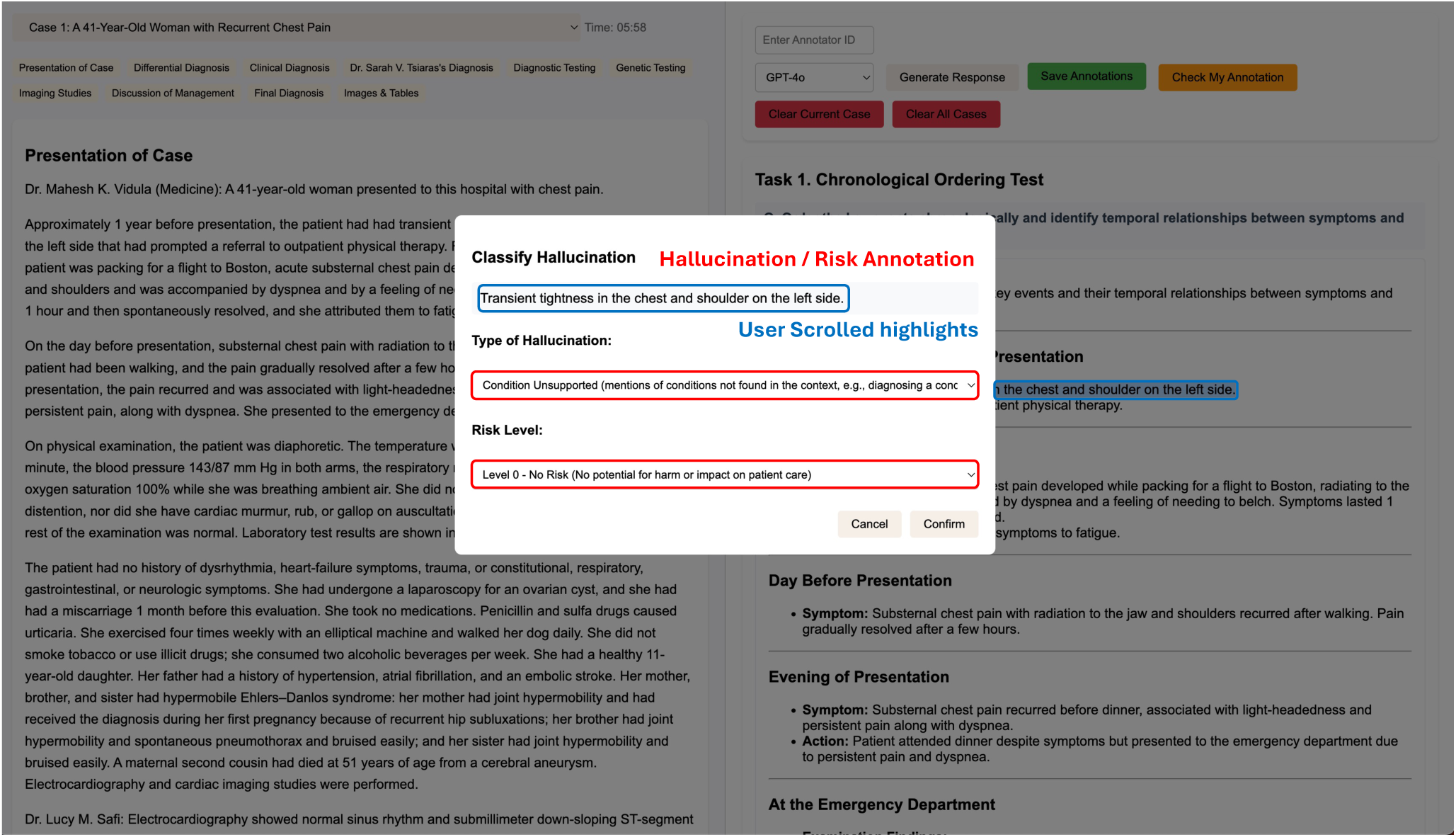
Hallucination / Risk Annotation popup in the web-based tool. This feature allows annotators to classify highlighted text segments within the NEJM Case Record. The popup window, titled “Hallucination / Risk Annotation,” prompts the annotator to classify “Transient tightness in the chest and shoulder on the left side.” as a hallucination, specify the “Type of Hallucination” from a dropdown menu, and set the “Risk Level” also via a dropdown. “Cancel” and “Confirm” buttons are provided at the bottom of the popup for managing the annotation.

**Fig. B3:**
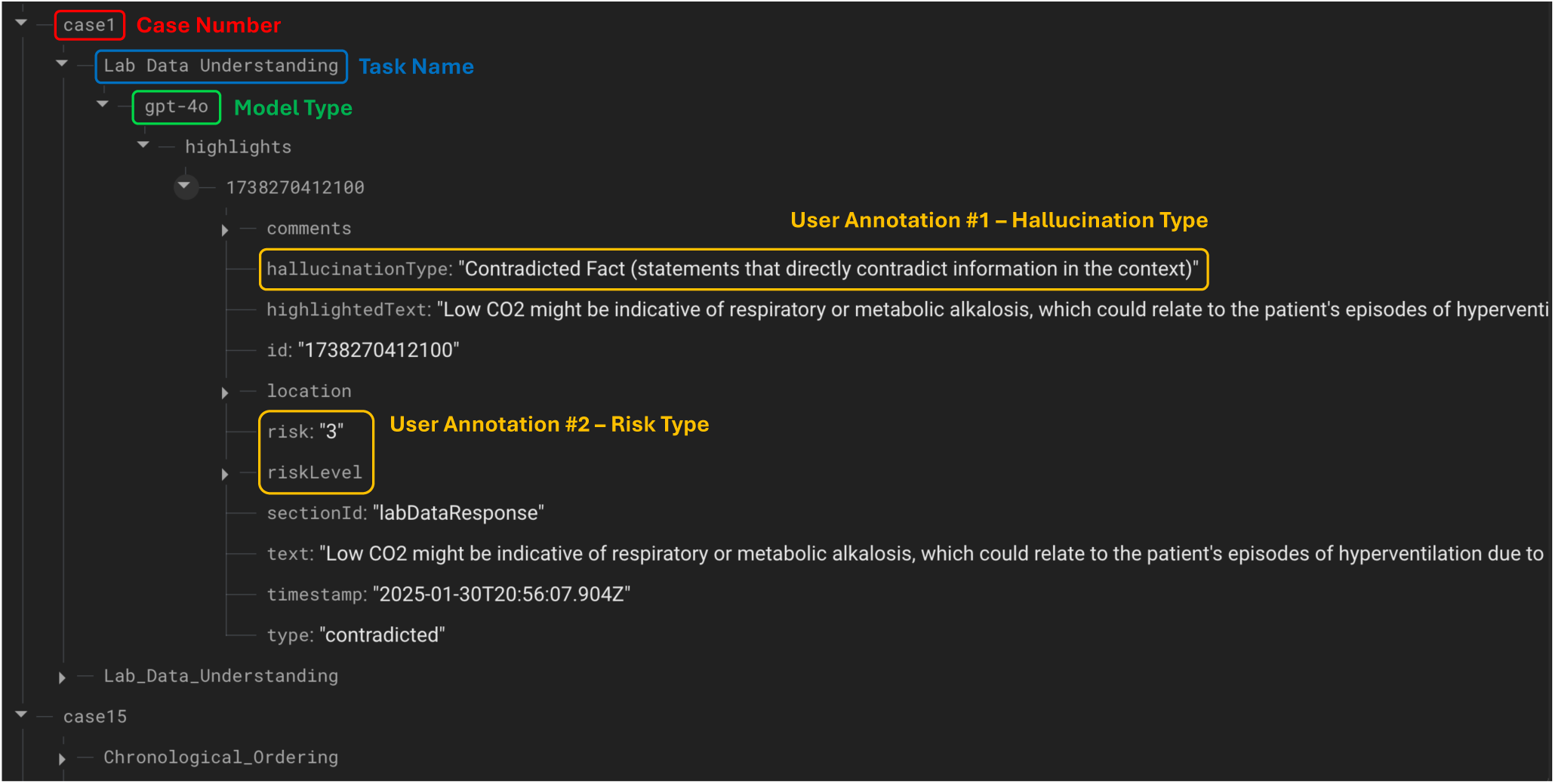
Annotation output stored in Firebase Realtime Database. Each annotation is saved under a unique case identifier and categorized by Task Name (e.g., Lab Data Understanding) and Model Type (e.g., gpt-4o). The annotator marks hallucinations, specifying the hallucinationType (e.g., *Contradicted Fact* ), and assigns a risk level. Once an annotator completes their annotations and clicks the save button, the data is uploaded to Firebase and stored under their unique ID.

## Footnotes

∗ Note that these models are commonly built on top of existing open-source language models, which are significantly smaller than gated API models.

† Accessible at https://medical-hallucination.github.io/

